# Combining blood biochemistry and glycan ages for an even more prognostic biological age

**DOI:** 10.1101/2025.09.14.25335700

**Authors:** Azra Frkatović-Hodžić, Jordan Bortz, Andrea Guarglia, Erin Macdonald Dunlop, Anika Mijakovac, Nina Šimunić Briški, Peter Ward, Ozren Polašek, Gordan Lauc, Michael Geer, James F Wilson, Peter K Joshi

## Abstract

Glycan- and blood marker–based biological ages are prognostic of health and survival. However, glycans and haematology measures (as opposed to blood biochemistry) are rarely available together in outcome-based studies. Consequently, no dataset available to us could unify these biological age estimates.

We therefore developed a new measure, *PlasmaAge*, using only biochemistry markers measured in ORCADES but trained within UK Biobank. In ORCADES, *PlasmaAge*, a glycan-based age, and their composite all predicted mortality, improving AUC over chronological age alone by 0.026, 0.032, and 0.041 respectively. We confirmed these effects on mortality in Croatian cohorts.

*PlasmaAge* provides a new estimate of biological ageing for studies where only biochemistry data are available. Moreover, we show that *PlasmaAge* and glycan-based age can be combined into a more prognostic composite biological age, suitable for use in both research and commercial settings.

## Introduction

Biological age (BA), the notion that functional age develops similarly, but not identically to chronological age (chronAge), was first postulated by Baker and Sprott (Baker and Sprott 1988). Interest in BA is increasing both as a predictive tool and as a target for ageing interventions and has led to the discovery of numerous biomarker based BA estimates.

There are two general approaches to the development of a biomarker based BA. The first, as exemplified by Hannum’s (Hannum et al. 2013) and Horvath’s (Horvath 2013) epigenetic clocks and Levine’s application of the Klemera and Doubal method (Levine 2013; Klemera and Doubal 2006) is to identify many age related markers and then use these to predict chronAge, the predictions themselves being defined as the subjects’ BAs. This can be done for a wide range of biomarkers in different domains, or by combining markers across domains. Essentially, this approach recognises that people in the population with their set of biomarker measurements are on average chronAge x, so postulates x is the subject’s BA. The approach is intuitive, especially if using a limited number or one biomarker, even if counter intuitively, the average person chronologically x aged may not have a BA of x (which we discuss further later). Furthermore, as the number of biomarkers increases, and with a sufficiently large training set, biological age converges to chronAge (Macdonald-Dunlop et al. 2022). Whilst a very accurate chronAge model is potentially of interest to understand the evolution of markers with chronAge, and might be of use, perhaps forensically, where chronAge is not easy to establish more directly, it (by definition) reveals little about functional age as distinct from chronAge. The second approach overcomes this by creating biological age models based on function, not chronAge as an outcome. A natural category of function is an age-related outcome, such as susceptibility to death or disease. Such outcome based clocks however require large datasets with an outcome tracked over time, to perform training, for example Liu et al’s phenoAge (Liu et al. 2018) and GrimAge (Lu et al. 2019). Such BA estimates have two advantages: first, they are by design optimal predictors of the (outcome-specific) functional age and second, they present a natural target for the purpose of intervention, in that they are predictive of better outcomes (recognising that confounding and reverse causality may mean that the predictor is not causal to the outcome). Further, if well calibrated such models can measure biological age acceleration (BAA), the excess (or deficit) of functional age over chronAge, with a mean of zero for subjects of any given chronAge. On the other hand, chronAge-based clocks are often more feasible to train (as the outcome - age at participation - is easily gathered) and the residuals of such models trained with an intermediate number of markers have been shown to be functionally meaningful, in particular predictive of mortality and disease (Marioni et al. 2015). The choice of a chronAge, or outcome-trained BA estimates is thus often pragmatic, with the proof of the pudding being in its purpose and utility.

Blood based clinical measures have often been used to develop estimates of mortality outcome-based biological age, for example phenoAge and Bortz BioAge (Liu et al. 2018; Bortz et al. 2023).

Conversely a model trained on chronAge enabled the development of a glycan clock of biological age (Krištić et al. 2014) – GlycansAge based on the measurement of glycans attached to immunoglobulin G (IgG). IgG is the most abundant antibody in blood, and holds a central role in the human immune system. The fragment crystallizable (Fc) region of IgG contains a conserved N-glycosylation site that predominately harbours different biantennary complex-type N-glycans (Pučić et al. 2011). They regulate the effector function of IgG by fine-tuning the interaction of the Fc region with various components of the immune system. Thus, the addition of different glycans can act as a molecular switch between pro- and anti-inflammatory immune responses (Krištić and Lauc 2024). Since chronic inflammation was formally recognized as a hallmark of ageing by Lopez-Otin and colleagues in 2023 (López-Otín et al. 2023), IgG glycans captured the attention of the ageing and longevity field. The intimate association of chronic inflammation with all other ageing hallmarks (López-Otín et al. 2023; Baechle et al. 2023) allows IgG glycans to measure biological ageing on the systemic level. Furthermore numerous studies have demonstrated that IgG glycans are exceptionally stable under homeostatic conditions but change when homeostasis is disturbed (e.g. ageing, disease, hormone fluctuations, environmental stimuli), as reviewed by Shkunnikova et al.(Shkunnikova et al. 2023) and Krištić and Lauc (Krištić and Lauc 2024). Moreover, they are responsive to different lifestyle and medical interventions (Shkunnikova et al. 2023).

However, in the absence of empirical evidence, it is, as yet, unclear if GlycansAge and blood BAs (such as phenoAge) are measuring the same or different aspects of ageing and such knowledge could inform creation of a composite, more prognostic BA, and also indicate to subjects which dimensions of ageing they have been subject to. Research dissecting these issues is difficult as few sufficiently large datasets are available where GlycansAge and a blood BA can be measured simultaneously, alongside an age-related outcome, such as death. For example, the ORCADES study of an archipelago in the north of Scotland,(McQuillan et al. 2008) has blood plasma-based measures, along with IgG glycans and has 13 years of follow-up for survival with ∼300 deaths. However, existing blood based BAs, for example phenoAge and BortzBloodAge have often used markers only present in whole blood (such as haematology measures, like red cell distribution width), making it impossible to measure those BAs in ORCADES, or other research cohorts where plasma rather than whole blood has been used for assays. On the other hand, UK Biobank (UKBB) (Sudlow et al. 2015) has extensive measures of biomarkers from blood alongside follow up for mortality, but, as yet, no IgG glycans assays. So, using these datasets, it is not feasible to address the question of the similarities or differences in GlycansAge and those blood BAs.

However, using UKBB it is possible to develop mortality outcome-based models of biological age using any set of markers desired, for practical application elsewhere. Importantly, the number of UKBB participants with blood data (>360,000) and the duration of follow up, means development of these models is typically well powered. It is therefore feasible to create new blood biomarker BA models in UKBB, targeted for use in other cohorts with only particular assays available.

Thus we reasoned, we could create a new blood-based biological age, plasmaAge, using only blood biochemistry assays available in ORCADES, which would have utility to the research community whose cohorts have only retained blood plasma, and would also enable us to study the separate and joint effects of PlasmaAge and GlycansAge, enabling a more holistic and/or accurate measurement of patients’ BA and, importantly, its dissection into component parts, guiding understanding and interventions. We then sought corroboration of our findings in Croatian island cohorts Vis and Korčula, where blood plasma measures and GlycansAge are also available.

## Results

### Development of PlasmaAge in UKBB

For our development of PlasmaAge in UKBB, 499652 subjects were available for analysis, of whom 366654 had complete data for our purposes. 54% were female, and 22,250 (6.1%) died. The mean follow-up time was 11.6 years, with a maximum of 14.0 years. Summary statistics for the UKBB population characteristics and the blood assays used can be found in Supplementary Table 1, and the distributions are depicted in Supplementary Figure 1a and 1b.

Using Cox proportional hazard (Cox PX) modelling (Cox 12 2018), we fitted a null model (age only) for survival, which showed log_e_ HR effect (SE) for age of 0.101 (0.0011) years (p<2e-16). The model concordance was 0.6956. In subsequent modelling, following previous work by us (Bortz et al. 2023), log_e_ Cox PH ratios were converted into units of age in year - to correspond to age accelerations by scaling, as described in Methods: a log_e_ HR of 0.1 corresponded to a 0.99 years of age acceleration.

The PlasmaAge model, which had 15 potential markers (listed in Supplementary Table 3), plus age, potentially available, and used 12 markers after dropping statistically non-significant calcium, cholesterol and triglycerides, obtained a concordance of 0.7245 (an increase of 0.0291 over chronAge alone). The model coefficients are shown in Table 1. As well as looking at coefficients on the equivalent years of age per unit marker scale, we looked at them on the per standard deviation scale, comparing on the per SD scale, the multivariate (MV) coefficient with that for age and single marker (univariate - i.e. a model using the variable alone, not even including age as an effect). Age had the largest effect on the standard deviation scale for both MV/UV models (7.40/8.09 years/SD), whilst glucose had the smallest absolute MV effect (0.14 years/SD). However, in a univariate model, urea had a much larger effect, (1.65 years/SD), whereas log_e_ alanine aminotransferase had the smallest absolute univariate effect (0.305 years/SD), but an absolutely larger and negative effect (−1.37 years/SD) in the multivariate model. The multivariate and univariate effect sizes (on the SD scale) are compared in Figure 1

**Table 1:** PlasmaAge model developed in UKBB.

| Marker | Coefficient | Standard Error | P Value | Coefficient per SD | Univ Effect per SD |
| --- | --- | --- | --- | --- | --- |
| <chr> | <dbl> | <dbl> | <chr> | <dbl> | <dbl> |
| age | 0.91 | 0.01 | 0.0e+00 | 7.40 | 8.09 |
| LDL | -4.85 | 0.31 | 7.9e-54 | -4.15 | -1.76 |
| APOB | 12.51 | 1.13 | 2.2e-28 | 2.96 | -1.03 |
| GGT | 0.06 | 0.00 | 9.4e-195 | 1.76 | 1.84 |
| ln_CRP | 1.71 | 0.07 | 1.7e-133 | 1.75 | 3.05 |
| APOA1 | -5.61 | 0.66 | 1.5e-17 | -1.50 | -1.51 |
| ln_ALT | -3.15 | 0.18 | 7.6e-66 | -1.37 | 0.31 |
| CREAT | 0.08 | 0.00 | 1.5e-55 | 1.11 | 1.96 |
| HBA1C | 0.20 | 0.01 | 8.1e-53 | 1.01 | 2.76 |
| HDL | 1.87 | 0.50 | 2.0e-04 | 0.69 | -1.99 |
| ALB | -0.26 | 0.03 | 2.1e-20 | -0.66 | -2.21 |
| UREA | -0.50 | 0.06 | 1.1e-18 | -0.64 | 1.65 |
| GLU | 0.18 | 0.08 | 2.2e-02 | 0.14 | 1.71 |

**Figure 1:**
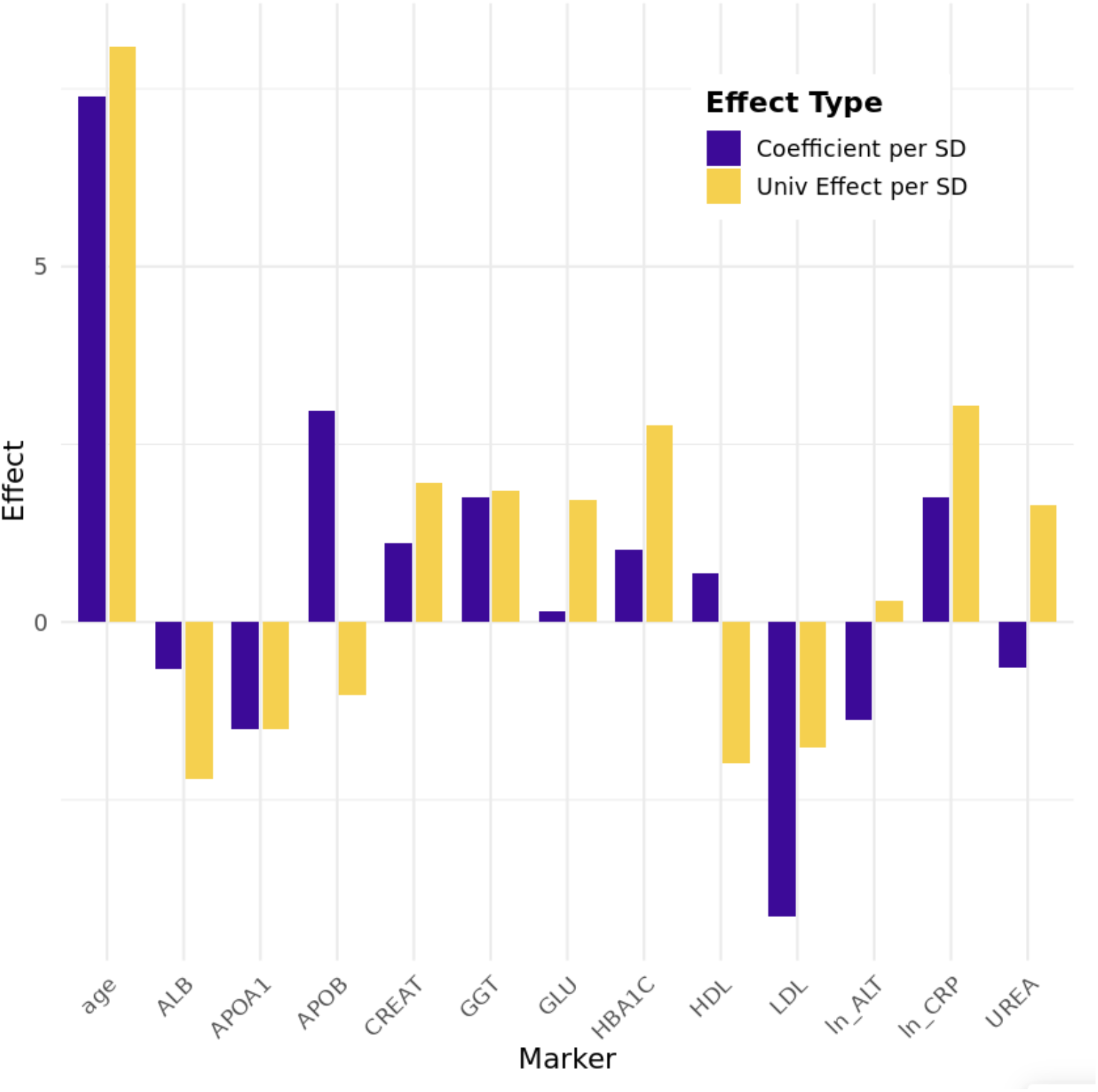
The effect of individual markers on BA in UK Biobank. Marker: the assay code The code and assay description and units are listed in Supplementary Table 2. Ln_: after log_e_ transformation. Coefficient per SD - change in age per one standard deviation of the predictor in the multivariate model. Univ Effect per SD - change in age per one standard deviation in univariate model.

We would later wish to test whether our findings for PlasmaAge were replicated in an independent dataset - using the Croatian island cohorts Korčula and Vis. However, not all PlasmaAge markers are available there and indeed the two cohorts had two different sets of measurements. We made the choice to develop cohort specific plasma ages (in UKBB), rather than reduce our marker set to the smaller common set, to provide the maximum predictive power in each cohort, given the limited size of each dataset. PlasmaAgeKorcula and PlasmaAgeVis models, were derived in exactly the same way, with the markers available (shown in Supplementary Table 3). The results are shown in Supplementary Table 4.

Staying with UKBB, we next compared the performance of this model with that of phenoAge, for reference. We calculated phenoAge directly, and also calculated a UKBB optimised set of coefficients using the same biomarkers (phenoAgeRecalibrated) in case population differences meant that the phenoAge model trained in NHANES could be improved for use within UKBB. As previously stated, age alone gave a concordance of 0.6956, PlasmaAge, a concordance of 0.7257, and phenoAge had a concordance of 0.7275. PlasmaAge thus achieved 94% of phenoAge’s whole blood BA predictive value (concordance over and above that of age alone), with results depicted in Figure 2. PhenoAgeRecalibrated gave a concordance of 0.7327, a 16% increase in added predictive power from standard phenoAge, thus plasmaAge achieved 81% of the added predictive power PhenoAgeRecalibrated. The performances of the plasmaAge, phenoAge and PhenoAgeRecalibrated in excess of age alone, expressed as a percentage of the increase achievable by a perfect predictor in excess of that for age alone were 9.9%, 10.5% and 12.2% respectively.

**Figure 2:**
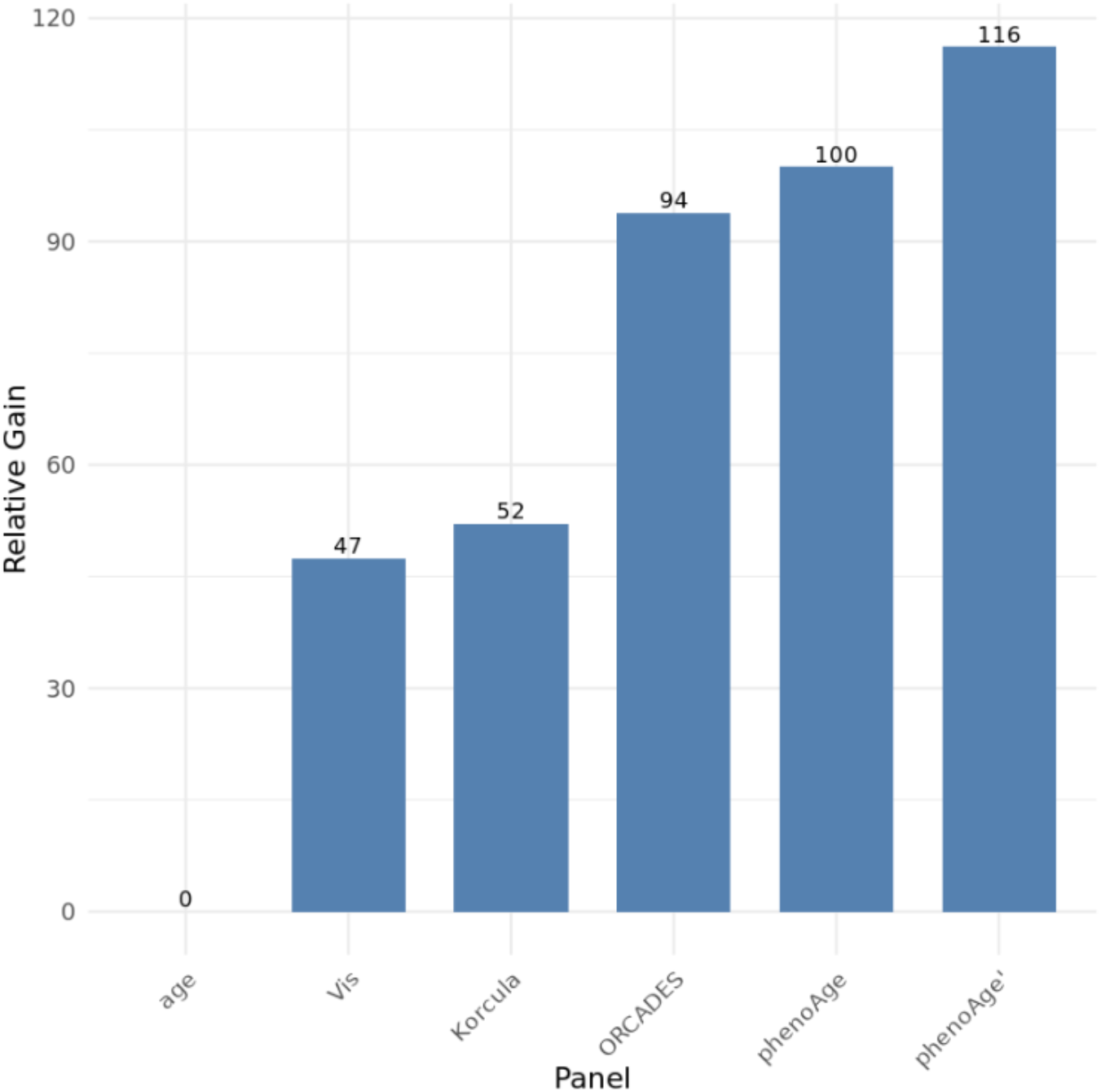
In UKBB, blood biochemistry markers can achieve a similar gain in survival concordance (over age alone) to phenoAge’s haematology and biochemistry based panel (pre and post recalibration of phenoAge for UKBB tailored effect sizes) Panel: the set of markers included for each analysis in UKBB of the different blood based BAs (the Vis/Korcula/ORCADES panels represent the (plasma based) biochemistry marker sets available in that cohort (see Supplementary Table 3) - but this analysis is in UKBB using that marker set) Relative Gain - the survival concordance achieved in UKBB by the marker set in excess of that achieved by a chronological age in the same sample set, expressed as a percentage of that achieved by phenoAge. phenoAge’: model based on the marker components of phenoAge, but with coefficients recalculated from their survival model in UKBB.

### Calculation of PlasmaAge and GlycansAge in ORCADES

We now had 3 PlasmaAge models (PlasmaAge, PlasmaAgeVis, PlasmaAgeKorcula), trained in UKBB which required only markers available in the cohort concerned and set to test their prediction power in the respective cohorts.

Of the 2,314 participants in the ORCADES cohort, 2,009 had complete basic phenotypic information recorded, i.e. age, gender, and survival status. Among these, 941 participants had the complete plasma and glycan information needed. Analysis was restricted to those aged between 40 and 70 at outset: the choice determined specifically by the age at outset range in which the UKBB training had been performed, but also to avoid using a very large chronAge range, whilst capturing a reasonable number of deaths. With too large a chronAge range, we found that chronAge explained too much mortality risk, leaving little room for other predictors, whereas if the age range was too narrow there were insufficient participants. Of the 595 participants in the target age range, with complete data, 0 were removed due to outlying age accelerations (AAs), leaving 595 participants to include in the final analysis. Of these 595 participants, 326 were female, and 76 had died, as displayed in Supplementary Figure 2 These are the subjects taken forward for analysis.

PlasmaAge and standard glycans age (GlycansAge), was calculated for all participants. We found that GlycansAge was an unbiased predictor of ChronAge (Supplementary Figure 3). GlycanAA correlated nevertheless with chronAge, as illustrated in Supplementary Figure 4. That correlation of GlycanAA with chronAge was undesirable for our purposes for two reasons. Firstly in a multivariate model, cross-talk between the two correlated features will occur, inhibiting interpretation, second, for individual reporting of AA, a statement to a 25 year old, that your GlycansAge is ∼38 but that’s true for the average 25 year old is confusing. The subject will usually want to know how their glycans profile compares with an average person their age, and presume mean AA is zero at their age.

We therefore decorrelated GlycanAA from chronAge by calculating a linear model, for participants age 40-70, of GlycanAA with chronAge and used those model residuals to define a revised Glycans age acceleration (GlycanA’A), with consequent decorrelated Glycans age (GlycanA’ = GlycanA’A+chronAge). *GlycansAge*′/*GlycanA*′*A* = −18.01 + *age* × 0.338 + *GlycansAge*/*GlycanAA* × 1.

Now, participants at any given age typically have on average a GlycansAge’ equal to their chronAge, as illustrated in Supplementary Figure 5.

We next examined the relationship between plasmaAA and glycanA’A, finding they were fairly independent (r=0.10), as illustrated in Figure 3.

**Figure 3:**
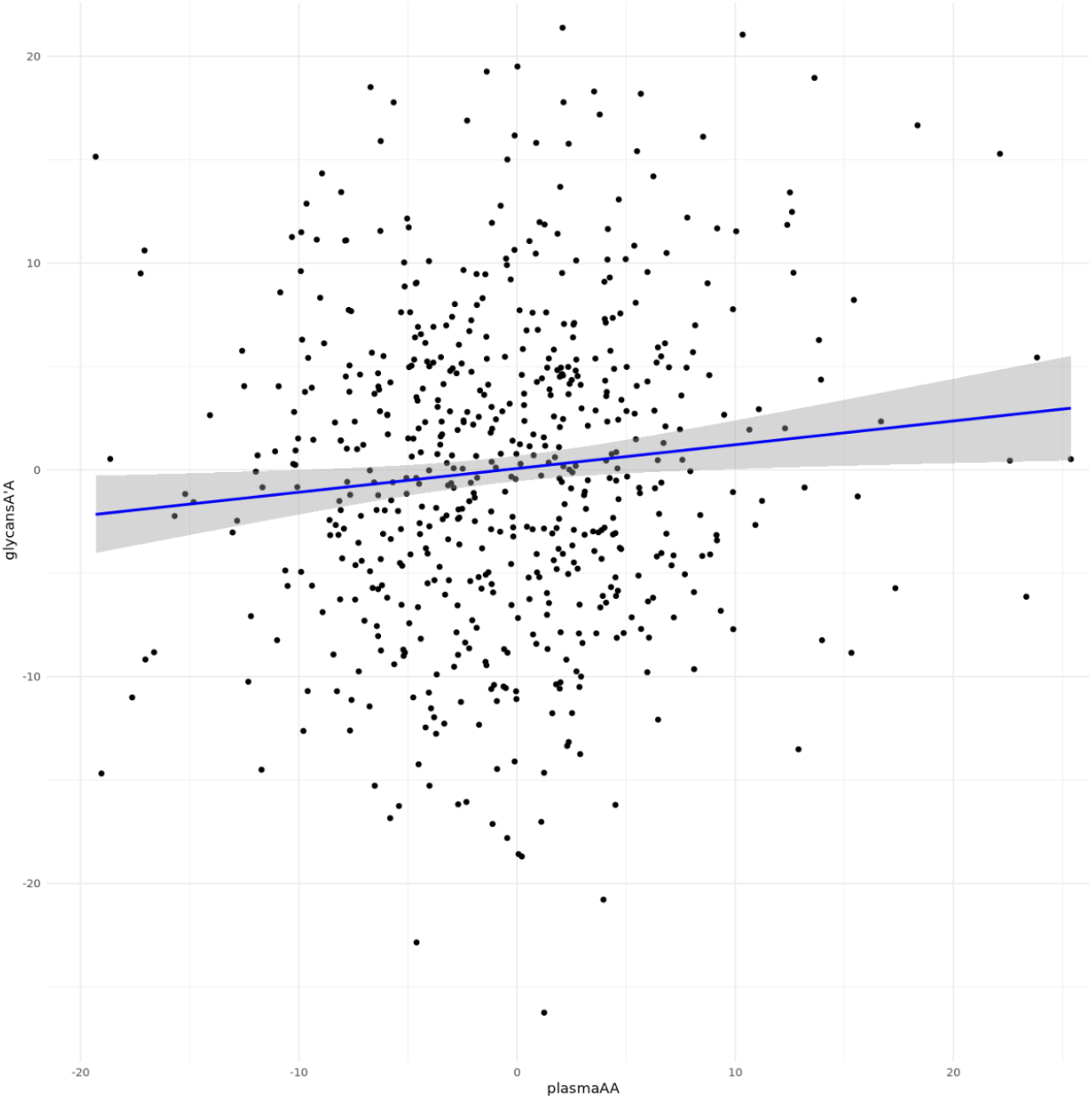
PlasmaAge and GlycansAge’ accelerations appear to track relatively independent aspects of aging in ORCADES. PlasmaAA: PlasmaAge acceleration in years,GycansA’A: chronAge decorrelated GlycansAge acceleration in years.

### Associations with mortality of PlasmaAge and GlycansAge in ORCADES

Next, we analysed the multivariate association between alive/dead status, with chronAge, and age accelerations, using a logistic regression model estimating log_e_ odds ratios for the event occurring, and calculating the AUC for the predictions.

In all models, associations of death with both age and each biological age acceleration were always statistically significant (p<0.001, one sided test), coefficients are shown in Table 6. Examining one AA at a time, we found that the coefficient was 0.73/0.58 years per year of Plasma/GlycansAge’ acceleration, noting that in UKBiobank for the same logistic regression model, the coefficient for plasmaAge was 1.02. The joint Plasma and GlycansAge’ accelerations model gave similar coefficients (0.65/0.51), to the single BAA analyses.

We next considered the predictive power of these models, as determined by AUC. The base model (age only) had an AUC of 0.734, the plasmaAA model, glycansAA model, and joint models achieved 0.028, 0.032 and 0.043 increases in AUC above that, as shown in Figure 4.

**Figure 4:**
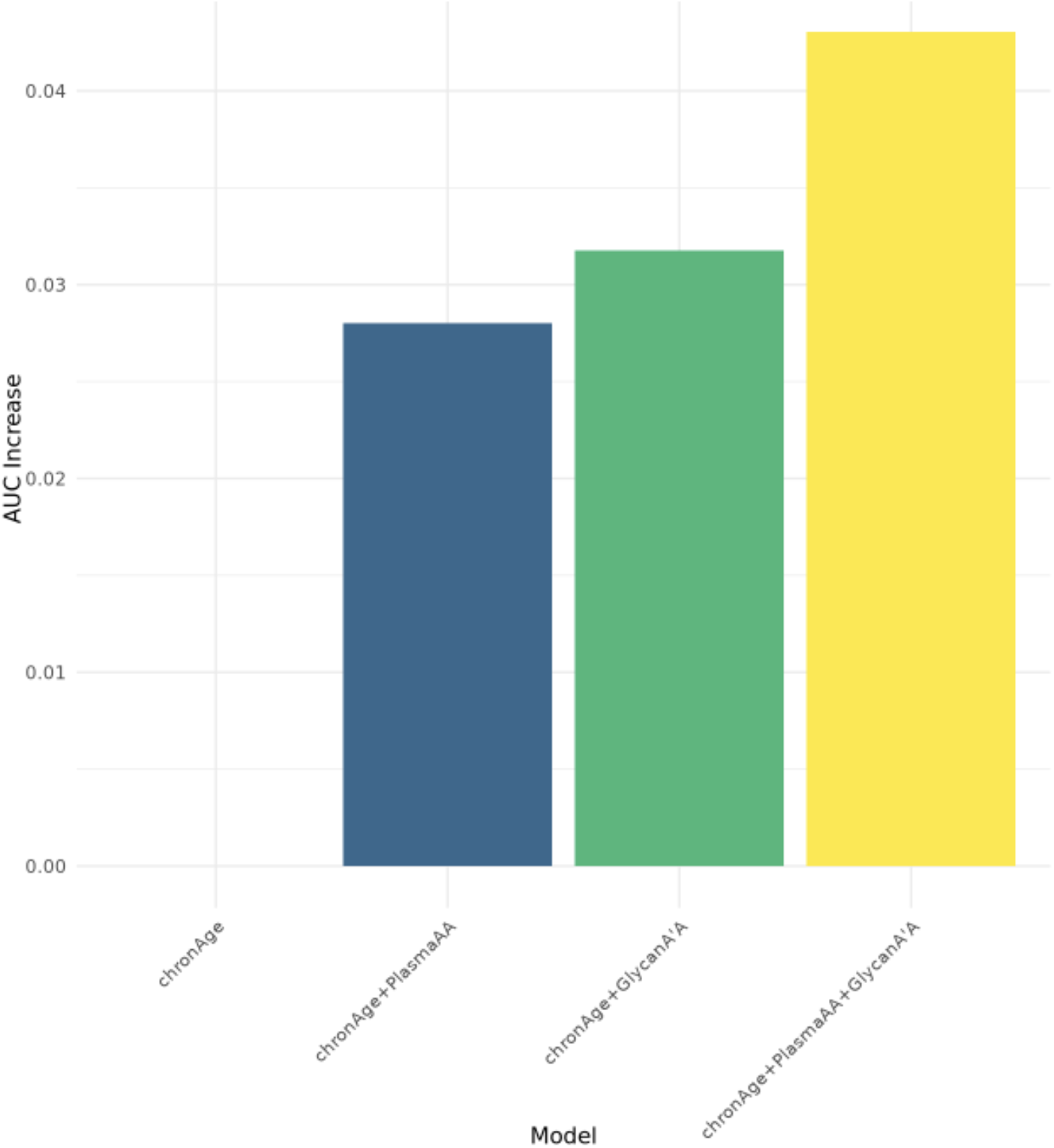
A composite model using GlycanA’A and PlasmaAgeA together achieves greater predictive power for mortality than each predictor alone. AUC increase: the AUC for the model over that for chronAge alone. Model: the effects fitted (chronAge+BAAs) fitted in the model.

Reviewing the chronAge + GlycanA’A column of Table 2, where 1 year of Glycan A’A acceleration gives rise to 0.5837 years of equivalent mortality increase, alongside the decorrelation calculation derived earlier, suggested a further GlycansAge recalibration - to bring GlycanAA to a scale of chronological age years of mortality from a scale of glycan years.

**Table 2:** Composite Biological Age Acceleration is a weighted sum of Plasma and Glycans Age accelerations in ORCADES.

| Effect | Model |  |  |  |
| --- | --- | --- | --- | --- |
|  | chronAge | chronAge + PlasmaAA | chronAge + GlycanA'A | chronAge + PlasmaAA + GlycanA'A |
| chronAge | 1 (-) | 0.99 (0.17) | 1.08 (0.18) | 1.06 (0.18) |
| GlycanA'A | - | - | 0.58 (0.16) | 0.51 (0.18) |
| PlasmaAA | - | 0.73 (0.18) | - | 0.65 (0.18) |

*Effect: The coefficients (SE) for the logistic model of death, re-expressed in equivalent years of age. Each model is represented by a separate column and its heading indicates the features included. The resultant value is the estimated (composite) biological age acceleration (i.e. the mortality rate increase in terms of equivalent years of chronAge) due to chron Age or BAA. Composite BA acceleration excess above average of each marker is simply the sum of the products of the observed values of each marker and the coefficient in the column. All effects were statistically significant ( $p < 0.001$ one-sided test).*

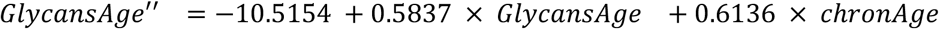

Finally, we sought to confirm our findings across the Croatian cohorts, where power was more limited, As previously noted there were 1,417 participants (Vis n=533; Korcula1 n=318; Korcula3 n=566), of whom 173 (Vis n=123; Korcula1 n=26; Korcula3 n=24) were dead. Within each cohort, we constructed the same logistic regression models and meta-analysed the effects sizes observed (noting that effect sizes for plasma{Cohort}AA models might be expected to be different, as different markers were used in Korčula and Vis, but understanding on the other hand that the calibration in UKBB had been in (mortality) equivalent years of age - suggesting similar effect sizes might be observed). The meta-analysis for Croatian cohorts showed wide confidence intervals for individual sub-cohorts, but revealed statistical significance (p<0.05, one sided t-test:H1 effect>0), for all the Croatian meta-analysed coefficient estimates (Table 3). Forest plots representing meta-analysis are shown in Supplementary Figure 7. ROC curves and graphs illustrating AUC values across different models for mortality prediction are shown in Supplementary Figure 8

**Table 3:** Composite Biological Age is a weighted sum of PlasmaAge and GlycansAge accelerations: replication in a meta-analysis of Croatian cohorts.

| Coefficient | chronAge | chronAge + PlasmaAA | chronAge + GlycansA'A | chronAge + PlasmaAA + GlycanA'A |
| --- | --- | --- | --- | --- |
| chronAge | 1 (-) | 1.01 (0.11) | 1.02 (0.11) | 1.03 (0.11) |
| GlycanA'A | - | - | 0.18 (0.12) | 0.15 (0.12) |
| PlasmaAA | - | 0.52 (0.209) | - | 0.50 (0.20) |

*Effect: The coefficients (SE) for the logistic model of death expressed in equivalent years of age. Each model is represented by a separate column and its heading indicates the features included). The resultant value is the estimated (composite) biological age acceleration (i.e. the the mortality rate increase in terms of equivalent years of chronAge) due to chron Age or BAA. Composite BA under each model (i.e. column) is simply the sum of the products of the observed values of each effect (composite) marker and the coefficient.*

## Discussion

We developed a new model of biological age, plasmaAge, in UK Biobank, whose acceleration, in conjunction with age, gave a survival concordance of 0.7296, compared to age alone of 0.7068. PlasmaAgeA achieved 90% of the increase in predictive power of phenoAgeA and 75% of the increase that a UKBB optimised model, using the phenoAge markers achieved. Apolipoprotein B had the largest biological age increasing effect (coefficient per biomarker SD), whilst LDL cholesterol had the most decreasing effect. We then analysed mortality using age and sex alongside GlycanA’A and PlasmaAA separately and then jointly in the independent Scottish cohort ORCADES and obtained AUCs for mortality of 0.026, 0.032 and 0.041 respectively in excess of that for chronAge. We derived a recalibration of GlycansAge (GlycansAge’’), which had two desirable properties: mean GlycanA’’A at each age was near zero and one year’s GlycanA’’A increases expected mortality by one year. We also found weak (r=0.1) correlation between GlycanA’A and PlasmaAA. PlasmaAA/glycanA’A had coefficients (i.e. the mortality effect equivalent to an increase in age of one year per year of AA) of 0.71/0.58 and 0.63/0.51 in single and joint models respectively.

This essentially compares subjects of the same age - with a mean near zero at each age. We found that participants with lower chronAges needed a downward adjustment, whilst those with higher chronAges needed an upwards adjustment: specifically

PlasmaAge is a new measure of biological age that can be used in research cohorts who have retained and analysed plasma rather than whole blood and thus have only biochemistry rather than haematology measures as well. It achieves reasonable predictive power in UKBiobank, where it was trained, and ORCADES where it was tested.

We suggest care must be taken in interpreting the coefficients of plasmaAge. Although the model as a whole results in an optimised mortality predictor, covariance amongst predictor coefficients means that interpretation of a predictor in isolation, may be misleading, as an individually apparently beneficial predictor may correlate positively with one or many deleterious predictors. Secondly, our study finds observational associations, and even greater care is needed before inferring causation. Our finding that higher LDL cholesterol is associated with reduced mortality is a case in point. The effect is reduced in a bivariate (age + LDL) setting, highlighting the issue of covariance with other markers, nonetheless higher LDL is still associated with lower mortality. This is inconsistent with findings that statins, which reduce LDL, lower cardiovascular disease risk (Mihaylova et al. 2024), nonetheless our finding is consistent with a previous observational (rather than RCT) work (Ravnskov et al. 2016). Our and Ravnskov’s findings, in our opinion, likely arise from confounding, reverse causality in a treated (with statins) population and perhaps, effects on other causes of mortality. Of course, none of this negates the predictive power of plasmaAge: it merely cautions against causal interpretations.

GlycansAge’A achieved a higher predictive value (AUC increase) than plasmaAge in our modelling. Considering the central position IgG glycans in immunity, as both biomarkers and functional mediators of inflammation, we hypothesize that GlycansAge reflects more nuanced aspects of immune function that are not entirely encompassed by other models, including PlasmaAge. The pivotal role of the immune system in ageing was highlighted by Prof. Claudio Franceschi when he introduced the concept of inflammageing. Inflammageing stands for the chronic, sterile, low-grade inflammation that develops with age as a consequence of the persistent, long-term activation of the immune system (C. Franceschi et al. 2000). Chronic inflammation was established as one of the 7 pillars of ageing along with adaptation to stress, epigenetics, macromolecular damage, metabolism, proteostasis, stem cells and regeneration (Kennedy et al. 2014). Markedly, all of the defined pillars are highly intertwined with all of them converging on inflammation as impairment of each one fuels inflammation, which in turn exacerbates the function of all others (Claudio Franceschi et al. 2018). Thus, a model that captures inflammageing, such as GlycansAge, can add a useful layer of information to existing data. Also, GlycansAge comes with one major advantage: the simplicity and affordability of the IgG glycomic analysis.(Trbojević-Akmačić et al. 2022). Moreover, GlycansAge requires only a few microliters of plasma to deliver robust results, eliminating the need for whole blood or large sample volumes, a common obstacle in many studies, and as we have shown here it is prognostic of future mortality.

By using the same age range in ORCADES as we had in UKBB, we expected the effect of a year of plasmaAgeA in ORCADES (0.71, SE 0.18 years of chronAge) to be similar to 1 and similar to that observed in UKBB (1.02, SE 0.02). Arguably it was, given a chance. However, the differing effects of chronAgeing, follow up periods and biomarkers in different populations may also be leading to this difference, as might the use of plasma rather than whole blood in the biochemistry assays. There is also a risk that cryptic incomplete follow up may be affecting our results, although that should affect chronAge effects too. On the other hand, we found evidence that one year of GlycansAge appears not to be associated with a mortality increase equivalent to one year of chronAge (effect=0.58 years of mortality age, SE 0.16). This is perhaps not surprising: GlycansAge has not been calibrated to have mortality effects on this scale, it has glycans effects of this scale. An acceleration in GlycansAge, thus appears to have a little over half the effect on overall health as a year of chronAge, whilst PlasmaAge plausibly has a full year’s effect (due to calibration). The issue here is one of scale, and of individual mortality rate prediction, not overall predictive power: glycans were more prognostic, based on the AUCs. We therefore propose that a mortality prediction-focused glycans age should take the raw glycans age acceleration and subject it to the age decorrelation shown and the subsequent contraction of 42%, derived above. A revised glycans age is then of course simply chronAge plus the age recalibrated age acceleration, improving interpretability, but not improving (or reducing) predictive power.

GlycansAge and PlasmaAge appear to be capturing distinct aspects of functional aging. They can be combined to form a composite measure which is even more predictive, and presumably more holistic than the separate measure alone. Moreover, where both are measured, they can provide guidance on an individual basis as to which dimension of BAA is the most impactful for the subject.

Our study confirms previous findings that BA, and in particular BAA, is measurable and meaningful and in particular can be predictive of survival and healthspan. Liu et al. (Liu et al. 2018) found similar associations of phenoAge with mortality in NHANES, findings previously confirmed by us in UKBB (Bortz et al. 2023), similarly, Marioni et al. (Bortz et al. 2023; Marioni et al. 2015) found accelerations in DNA methylation age were associated with mortality. Quantitative comparison of the results is not straightforward, due to the differences in dataset structure and no readily comparable summary statistics being available. We hope that more research in future will compare several clocks in the same dataset, but also suggest, where as will often be the case, this is not possible, that researchers report the prediction power (concordance or AUC) of chonAge alone against any model incorporating BA.

The strengths of our study were the large scale of UKBB enabling tight calibration of PlasmaAge, the availability of longitudinal prospective outcomes in the independent ORCADES cohort, alongside both IgG glycans and plasma assays, with sufficient power to achieve statistical significance in the mortality models, and estimate effect sizes. We also transparently measure the excess value of our biological ages over and above chronAge alone: without such knowledge users risk not knowing if biological age is simply an over complex way of calculating chronAge with noise.

Our study generalizability may be limited by the slightly unusual populations used, and the focus on age 40-70 at outset (albeit this is perhaps the age range where understanding ageing and potential interventions may be most valuable (Sudlow et al. 2015)). The relatively small scale of the studies also means that the coefficients derived have material uncertainty, but remain best estimates until more studies can be added. The heterogeneity of available markers between the studies used also hampered interpretation and meta-analysis.

Future work should include extending the number of studies measuring glycans age and indeed other biological ages, and, much more simply, calculating PlasmaAge in many more cohorts where the assay results needed are already available, and analysing the association between PlasmaAge and age related health outcomes. This might enable the investigation of the biological functions and outcome effects of the joint and independent aspects of GlycansA’’A and PlasmaAA, and this might in turn lead to investigations of the different causes of these forms of aging, guiding preventative and perhaps restorative interventions tailored to each subject’s aging profile.

PlasmaAge is a new measure of biological age, with potential widespread applicability in research cohorts and offers studies with only blood plasma markers the prospect of studying biological age, where they previously been precluded, due to not having all the markers required by slightly more accurate models requiring additional biomarkers - typically blood counts - to be measured. PlasmaAge and GlycansAge capture distinct dimensions of biological ageing and a composite PlasmaGlycansAge is an improved measure of ageing and health outlook.

## Online Methods

### COHORT descriptions

This study uses data from UK Biobank, a large, population-based prospective cohort that includes in-depth genetic and health information from over 500,000 individuals recruited between 2006 and 2010 across England, Scotland, and Wales (Sudlow et al. 2015). For this study, accessed data included recruitment date, age at recruitment, sex, date of death (if applicable), and blood biochemistry markers measured from samples collected at the time of recruitment.

### ORCADES

The Orkney Complex Disease Study (ORCADES) is a population-based cohort recruited in the isolated archipelago of the Orkney Isles in northern Scotland (McQuillan et al. 2008). 2078 participants aged 16-100 years were recruited between 2005 and 2011, most having three or four grandparents from Orkney, the remainder with two Orcadian grandparents. Fasting blood samples were collected and many health-related phenotypes and environmental exposures were measured in each individual, including extensive multi-omic characterisation. Multiple blood tubes were drawn after an overnight fast, including an EDTA tube, which was delivered within hours of venepuncture to the local NHS Orkney hospital laboratory. Clinical biochemistry measures were carried out at the Balfour Hospital using standard procedures on auto-analysers, after sample centrifugation.

Thirteen years of follow up is available via linked electronic health records, including linkage to the General Register Office for Scotland / National Records of Scotland death registry. The data linkage and access to NHS Scotland-originated data was approved by the Public Benefit and Privacy Panel for Health and Social Care (Ref 1718-0380).

### CROATIA cohorts

Croatian cohorts, Vis and Korčula, were collected as part of the “10001 Dalmatians” study of Croatian island isolates (Rudan et al. 2009). Participants in the Vis cohort (n=890) were sampled in 2003, while Korčula participants were sampled in two batches: Korcula-1 in 2007 (n=915) and Korcula-3 in 2013 (n=951).

Participants from the three cohorts were interviewed by trained surveyors using a comprehensive questionnaire. The questionnaire included information on personal characteristics (including name, date and place of birth, gender, marital status, education level, and occupation), health-related lifestyle factors (such as diet and smoking status), health complaints, medication use, and hospitalization history. Blood samples were collected in anticoagulant-containing tubes, immediately processed, and centrifuged to separate plasma, which was then stored at −70 °C for future analysis.

Death records for Croatian cohorts were accessed through Croatian national health registry in October 2022.

### UPLC measurements of IgG N-glycome

IgG N-glycan measurements for all four cohorts (Korcula1, Korcula3, Vis and ORCADES) were obtained using ultra-performance liquid chromatography (UPLC), as previously described (Pucić et al. 2011). Chromatograms obtained from the analysis were divided into 24 peaks (GP1– GP24), with glycan abundance expressed as a percentage of the total area. After preprocessing of the glycan data which included normalization and batch correction, three glycans were used in this study for calculation of GlycansAge: a nongalactosylated glycan (GP6) and two digalactosylated glycans (GP14 and GP15).

### Development of PlasmaAge in UK Biobank

We first investigated which UK Biobank blood biomarkers were available in ORCADES and Croatian cohorts. Supplementary Table 3 lists 19 UKBB blood markers (plus sex and chronAge) that are used in PhenoAge, or available in ORCADES (n=13). Of those present in ORCADES, 9 are present in Korcula-1 and Korcula-3, while 6 are present in Vis cohort. PhenoAge markers not available in ORCADES are mainly those haematology whole blood characteristics not available in plasma, whilst for Croatian cohorts some plasma/biochemistry markers have also not been assayed.

Of the 499,999 UK biobanks subjects in our dataset, 368,053 had complete data for chronAge, sex, survival and the 19 biomarkers used in PhenoAge, or available in ORCADES or Croatian cohorts. Survival was measured from the date of attending assessment centre until 27 Nov 2020 (reflecting the last date available at date of data download). These 368,053 were therefore taken forward for analysis, and comparisons.

Next, we visually examined the distributions of the biomarkers, and in particular focused on whether there was substantial right skew (Supplementary Fig. 1a and Fig. 1b). Alanine aminotransferase, alkaline phosphatase, C reactive protein, gamma glutamyl transferase and triglycerides all exhibited substantial right skew, and were transformed by taking log_e_, which did result in much more normally distributed values (Supplementary Fig. 1c and 1d).

For each set of available (or where appropriate log_e_ available) markers, we used Cox Proportional Hazard (PH) modelling (Cox 12 2018), predictors of mortality, in a model using those markers and chronAge with effectiveness of each model measured using Harrelson’s Concordance. For a given set of candidate markers, we fitted survival time from participation until study end (or death) and the presence or absence of death against the markers. If the resultant model showed that the marker was not statistically significant (in the presence of the other markers), it was excluded and the modelling rerun once, to give estimates of the log_e_ HR for each significant marker, in the multivariate model. To calculate PlasmaBA, we rescaled each effect. We calculated age_eff on log_e_ HR scale, the effect of chronAge on a model with only chronAge fitted. We then divided observed effects of all markers by age_eff. Thus the effect of 1 year of chronAge in the age only model was 1 year - i.e. we had a scale of BA in years of age, with the appropriate effect on mortality of one year of chronological aging. Confusingly, at first sight, this does not always lead to an effect of one year, per year of chronAge in the full biomarker models. This arises due to covariance between the (ageing) markers and chronAge. Biological Age acceleration (BAA), the excess of BA over chronAge, thus often includes a small negative term for chronAgeCA - and decorrelates BAA from chronAgeCA, as is desirable.

### Derivation of BAs and subsequent modelling of Mortality in ORCADES and Croatia

PlasmaAge BA and BAA were calculated using the models derived from UK Biobank, whilst phenoAge was calculated in accordance with Liu et al (Liu et al. 2018), and GlycansAge was calculated according to Krištić et al. (Krištić et al. 2014), using GP6, GP14 and GP15 glycan peaks. GPn represents n*th* peak in chromatogram expressed as proportion of total chromatogram area. Age model coefficients were recalculated in ORCADES to obtain cohort-specific coefficients, with separate models derived for males and females. Raw GlycansAge was calculated using the following formulas:

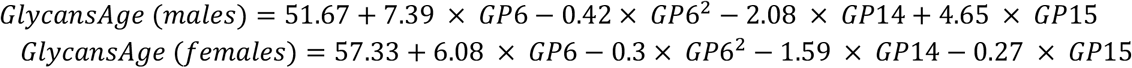

Base GlycanAA associated strongly negatively (r=-0.67) with chronAge. As our aim was to understand BA, over and above chronAge, this was undesirable. We therefore calculated a residualised (for age) GlycanAA under a linear model with GlycanAA as outcome and chronAge as predictor, which by design leaves a correlation of 0 between GlycanAA (the residuals) and chronAge, and provides a measure of glycans ageing for each individual, relative to people of their chronAge, with an expected mean of zero at each chronAge.

We wanted to exclude substantial outliers from subsequent analysis, whilst maximizing the amount of useful data available for analysis. In particular we didn’t want to drop a subject, if a relatively unimportant biomarker was not excessively out of range. The obvious way to assess importance was thus to consider the BA model coefficients, and thus we simply calculated BAA for each subject and excluded outliers where either age acceleration was in excess of 4 standard deviations from the mean.

To assess the effect on mortality in ORCADES and Croatia, because death as a binary outcome but not survival time data was available, a generalized linear model was fitted using a logistic regression approach, with a binomial error distribution and a logit link function, with dead as the binary outcome, and chronAge and (single or joint) BAAs as explanatory variables. Model coefficients thus represent the log-odds of the outcome per unit of explanatory variable. Again we rescaled these coefficients to the years of mortality age scale by dividing by the effect of chronAge in the model. R packages: survival, pROC and base glm where used to undertake the calculations.

## Data Availability

All data produced in the present study are available from UK Biobank or are available upon reasonable request to the authors

## Acknowledgements

We thank the UK Biobank Resource, approved under application 69634. We acknowledge funding from Humanity Inc, a company dedicated to measuring and improving biological age. The Orkney Complex Disease Study (ORCADES) was supported by the Chief Scientist Office of the Scottish Government (CZB/4/276, CZB/4/710), a Royal Society URF to J.F.W., the MRC Human Genetics Unit quinquennial programme “QTL in Health and Disease”, Arthritis Research UK and the European Union framework program 6 EUROSPAN project (contract no. LSHG-CT-2006-018947). We would like to acknowledge the invaluable contributions of the research nurses in Orkney, the administrative team in Edinburgh and the people of Orkney.

## Contributions

AFH, JB, PKJ. NSB, GL, PW, and MG participated in conceiving the study objectives and design. AFH, PKJ and JB carried out initial data preparation. AFH and PKJ performed the data analysis and model development. AFH and PKJ wrote the first draft of the manuscript. All authors reviewed and provided critical commentary on the manuscript. JFW provided the ORCADES data, OP provided the Croatia data. PW and GL provided funding for the project.

## Ethics declarations

### Competing interests

The authors declare the following competing interests: during preparation of this manuscript, PKJ and JB were paid consultants to Humanity Inc, a company focussed on measuring and developing interventions for Biological Age. MG and PW are founders of Humanity Inc and are employees and hold ordinary shares. PKJ, MG and PW are partly remunerated under a Humanity Inc share option scheme. PKJ is founder of Geromica, a consultancy providing advice on measurement of health and ageing. GL is the cofounder and CSO of GlycanAge Ltd. – the company that offers the glycan-based test for biological age. All other authors declare no competing interests.

### Ethical approval

This work used existing datasets (UK Biobank), for which ethical approval (including informed consent) had been obtained by UK Biobank for health investigation at the time of collection. This project was approved under UK biobank project 69634.

All participants in Croatian island cohorts signed an informed consent and study was approved by the Ethics Committee of the University of Split Medical School.

For ORCADES, all participants gave written informed consent and the study holds favourable opinions from Research Ethics Committees in Orkney, Aberdeen (North of Scotland REC), and South East Scotland REC, NHS Lothian (reference: 12/SS/0151). ORCADES is now part of Viking Genes (https://viking.ed.ac.uk/), under South East Scotland REC, NHS Lothian (reference 19/SS/0104; IRAS 264868).

## Notes

### Funding Statement

This study did not receive any funding.

### Author Declarations

This work used existing datasets (UK Biobank), for which ethical approval (including informed consent) had been obtained by UK Biobank for health investigation at the time of collection. This project was approved under UK biobank project 69634. All participants in Croatian island cohorts signed an informed consent and study was approved by the Ethics Committee of the University of Split Medical School. For ORCADES, all participants gave written informed consent and the study holds favourable opinions from Research Ethics Committees in Orkney, Aberdeen (North of Scotland REC), and South East Scotland REC, NHS Lothian (reference: 12/SS/0151). ORCADES is now part of Viking Genes (https://viking.ed.ac.uk/), under South East Scotland REC, NHS Lothian (reference 19/SS/0104; IRAS 264868).

## REFERENCES

Baechle, Jordan J., Nan Chen, Priya Makhijani, Shawn Winer, David Furman, and Daniel A. Winer. 2023. “Chronic Inflammation and the Hallmarks of Aging.” Molecular Metabolism 74 (101755): 101755.

Baker, G. T., 3rd, and R. L. Sprott. 1988. “Biomarkers of Aging.” Experimental Gerontology 23 (4-5): 223–39.

Bortz, Jordan, Andrea Guariglia, Lucija Klaric, David Tang, Peter Ward, Michael Geer, Marc Chadeau-Hyam, Dragana Vuckovic, and Peter K. Joshi. 2023. “Biological Age Estimation Using Circulating Blood Biomarkers.” Communications Biology 6 (1): 1089.

Cox, D. R. 12 2018. “Regression Models and Life-Tables.” Journal of the Royal Statistical Society 34 (2): 187–202.

Franceschi, C., M. Bonafè, S. Valensin, F. Olivieri, M. De Luca, E. Ottaviani, and G. De Benedictis. 2000. “Inflamm-Aging. An Evolutionary Perspective on Immunosenescence.” Annals of the New York Academy of Sciences 908 (1): 244–54.

Franceschi, Claudio, Paolo Garagnani, Paolo Parini, Cristina Giuliani, and Aurelia Santoro. 2018. “Inflammaging: A New Immune-Metabolic Viewpoint for Age-Related Diseases.” Nature Reviews. Endocrinology 14 (10): 576–90.

Hannum, Gregory, Justin Guinney, Ling Zhao, Li Zhang, Guy Hughes, Srinivas Sadda, Brandy Klotzle, et al. 2013. “Genome-Wide Methylation Profiles Reveal Quantitative Views of Human Aging Rates.” Molecular Cell 49 (2): 359–67.

Horvath, Steve. 2013. “DNA Methylation Age of Human Tissues and Cell Types.” Genome Biology 14 (10): R115.

Kennedy, Brian K., Shelley L. Berger, Anne Brunet, Judith Campisi, Ana Maria Cuervo, Elissa S. Epel, Claudio Franceschi, et al. 2014. “Geroscience: Linking Aging to Chronic Disease.” Cell 159 (4): 709–13.

Klemera, Petr, and Stanislav Doubal. 2006. “A New Approach to the Concept and Computation of Biological Age.” Mechanisms of Ageing and Development 127 (3): 240–48.

Krištić, Jasminka, and Gordan Lauc. 2024. “The Importance of IgG Glycosylation-What Did We Learn after Analyzing over 100,000 Individuals.” Immunological Reviews 328 (1): 143–70.

Krištić, Jasminka, Frano Vučković, Cristina Menni, Lucija Klarić, Toma Keser, Ivona Beceheli, Maja Pučić-Baković, et al. 2014. “Glycans Are a Novel Biomarker of Chronological and Biological Ages.” The Journals of Gerontology. Series A, Biological Sciences and Medical Sciences 69 (7): 779–89.

Levine, Morgan E. 2013. “Modeling the Rate of Senescence: Can Estimated Biological Age Predict Mortality More Accurately than Chronological Age?” The Journals of Gerontology. Series A, Biological Sciences and Medical Sciences 68 (6): 667–74.

Liu, Zuyun, Pei-Lun Kuo, Steve Horvath, Eileen Crimmins, Luigi Ferrucci, and Morgan Levine. 2018. “A New Aging Measure Captures Morbidity and Mortality Risk across Diverse Subpopulations from NHANES IV: A Cohort Study.” PLoS Medicine 15 (12): e1002718.

López-Otín, Carlos, Maria A. Blasco, Linda Partridge, Manuel Serrano, and Guido Kroemer. 2023. “Hallmarks of Aging: An Expanding Universe.” Cell 186 (2): 243–78.

Lu, Ake T., Austin Quach, James G. Wilson, Alex P. Reiner, Abraham Aviv, Kenneth Raj, Lifang Hou, et al. 2019. “DNA Methylation GrimAge Strongly Predicts Lifespan and Healthspan.” Aging 11 (2): 303–27.

Macdonald-Dunlop, E., N. Taba, L. Klarić, A. Frkatović, R. Walker, C. Hayward, T. Esko, et al. 2022. “A Catalogue of Omics Biological Ageing Clocks Reveals Substantial Commonality and Associations with Disease Risk.” Aging 14 (2). 10.18632/aging.203847.

Marioni, Riccardo E., Sonia Shah, Allan F. McRae, Brian H. Chen, Elena Colicino, Sarah E. Harris, Jude Gibson, et al. 2015. “DNA Methylation Age of Blood Predicts All-Cause Mortality in Later Life.” Genome Biology 16 (January):25.

McQuillan, Ruth, Anne-Louise Leutenegger, Rehab Abdel-Rahman, Christopher S. Franklin, Marijana Pericic, Lovorka Barac-Lauc, Nina Smolej-Narancic, et al. 2008. “Runs of Homozygosity in European Populations.” American Journal of Human Genetics 83 (3): 359–72.

Mihaylova, Borislava, Runguo Wu, Junwen Zhou, Claire Williams, Iryna Schlackow, Jonathan Emberson, Christina Reith, et al. 2024. “Lifetime Effects and Cost-Effectiveness of Statin Therapy for Older People in the United Kingdom: A Modelling Study.” Heart (British Cardiac Society) 110 (21): 1277–85.

Pucić, Maja, Ana Knezević, Jana Vidic, Barbara Adamczyk, Mislav Novokmet, Ozren Polasek, Olga Gornik, et al. 2011. “High Throughput Isolation and Glycosylation Analysis of IgG-Variability and Heritability of the IgG Glycome in Three Isolated Human Populations.” Molecular & Cellular Proteomics: MCP 10 (10): M111.010090.

Ravnskov, Uffe, David M. Diamond, Rokura Hama, Tomohito Hamazaki, Björn Hammarskjöld, Niamh Hynes, Malcolm Kendrick, et al. 2016. “Lack of an Association or an Inverse Association between Low-Density-Lipoprotein Cholesterol and Mortality in the Elderly: A Systematic Review.” BMJ Open 6 (6): e010401.

Rudan, Igor, Ana Marusić, Stipan Janković, Kresimir Rotim, Mladen Boban, Gordan Lauc, Ivica Grković, et al. 2009. “‘10001 Dalmatians:’ Croatia Launches Its National Biobank.” Croatian Medical Journal 50 (1): 4–6.

Shkunnikova, Sofia, Anika Mijakovac, Lucija Sironic, Maja Hanic, Gordan Lauc, and Marina Martinic Kavur. 2023. “IgG Glycans in Health and Disease: Prediction, Intervention, Prognosis, and Therapy.” Biotechnology Advances 67 (108169): 108169.

Sudlow, Cathie, John Gallacher, Naomi Allen, Valerie Beral, Paul Burton, John Danesh, Paul Downey, et al. 2015. “UK Biobank: An Open Access Resource for Identifying the Causes of a Wide Range of Complex Diseases of Middle and Old Age.” PLoS Medicine 12 (3): e1001779.

