## Supplementary material for "Combining blood biochemistry and glycan ages for an even more prognostic biological age": Supp Info

### Supplementary Tables and Figures

**Supplementary Table 1:** Summary statistics for personal characteristics and biomarkers of subjects with complete data in UK Biobank, used to create blood-based age models.

| marker | n | min | max | median | iqr | mean | sd | name | unit |
|---|---|---|---|---|---|---|---|---|---|
| <fct> | <dbl> | <dbl> | <dbl> | <dbl> | <dbl> | <dbl> | <dbl> | <chr> | <chr> |
| event | 366654 | 0.000 | 1.000 | 0.000 | 0.000 | 0.061 | 0.239 | Dead | 0=survived, 1 = Died |
| survival_yrs | 366654 | 0.011 | 14.008 | 11.852 | 1.430 | 11.650 | 1.547 | Survival time | years |
| age | 366654 | 37.000 | 73.000 | 58.000 | 13.000 | 56.528 | 8.091 | age | years |
| sex | 366654 | 0.000 | 1.000 | 0.000 | 1.000 | 0.460 | 0.498 | sex | 0=female, 1=male |
| ALT | 366654 | 3.010 | 94.450 | 20.020 | 11.660 | 22.851 | 11.318 | Alanine Aminotransferase | U/L |
| ALB | 366654 | 32.100 | 58.210 | 45.220 | 3.390 | 45.250 | 2.571 | Albumin | g/L |
| ALP | 366654 | 9.400 | 215.700 | 79.900 | 28.100 | 82.603 | 22.510 | Alkaline Phospatase | U/L |
| CHO | 366654 | 1.906 | 11.383 | 5.662 | 1.494 | 5.704 | 1.119 | Cholesterol | mmol/L |
| CREAT | 366654 | 10.800 | 165.000 | 70.500 | 19.400 | 71.998 | 14.417 | Creatinine | µmol/L |
| CRP | 366654 | 0.080 | 24.400 | 1.300 | 2.020 | 2.310 | 2.979 | High Sensitivity C-Reactive Protein (CRP) | mg/L |
| CYSC | 366654 | 0.295 | 1.789 | 0.885 | 0.174 | 0.902 | 0.144 | Cystatin – C | mg/L |
| GLU | 366654 | 0.995 | 11.339 | 4.919 | 0.698 | 5.030 | 0.810 | Glucose | mmol/L |
| HLRP | 366654 | 0.000 | 2.056 | 0.357 | 0.248 | 0.394 | 0.199 | High Light scatter Reticulocytes % | % |
| LYMP | 366654 | 0.000 | 66.300 | 28.620 | 9.520 | 28.945 | 7.260 | Lymphocyte percent | % |
| MCV | 366654 | 68.100 | 114.100 | 91.220 | 5.270 | 91.184 | 4.275 | Mean Corpuscular (erythrocyte) Volume | fL |
| MSCV | 366654 | 56.310 | 109.410 | 82.680 | 6.690 | 82.860 | 5.107 | Mean Sphered Cell Volume | fL |
| PCT | 366654 | 0.002 | 0.477 | 0.229 | 0.060 | 0.233 | 0.047 | Platelet crit | % |
| PDW | 366654 | 14.130 | 19.100 | 16.410 | 0.680 | 16.482 | 0.513 | Platelet Distribution Width | % |
| RDW | 366654 | 10.780 | 18.430 | 13.320 | 0.920 | 13.442 | 0.823 | Red Cell (erythrocyte volume) Distribution Width | % |
| WBC | 366654 | 0.000 | 17.450 | 6.630 | 2.180 | 6.841 | 1.730 | White Blood Cell or leukocyte count | x10^9 cells/L |
| HBA1C | 366654 | 15.000 | 70.000 | 35.200 | 5.000 | 35.663 | 5.063 | Glycated haemoglobin | mmol/mol |
| HDL | 366654 | 0.252 | 3.355 | 1.402 | 0.496 | 1.447 | 0.368 | High Density Lipoprotein (HDL) | mmol/L |
| LDL | 366654 | 0.806 | 7.667 | 3.535 | 1.163 | 3.575 | 0.856 | Low Density Lipoprotein (LDL) | mmol/L |
| TRIG | 366654 | 0.258 | 6.884 | 1.479 | 1.085 | 1.719 | 0.944 | Triglyceride | mmol/L |
| APOA1 | 366654 | 0.420 | 2.500 | 1.515 | 0.348 | 1.542 | 0.268 | Apolipoprotein A1 | g/L |
| APOB | 366654 | 0.400 | 2.000 | 1.019 | 0.317 | 1.034 | 0.237 | Apolipoprotein B | g/L |
| CA | 366654 | 1.914 | 2.850 | 2.376 | 0.117 | 2.380 | 0.092 | Calcium | mmol/L |
| GGT | 366654 | 5.000 | 247.900 | 26.100 | 21.600 | 34.680 | 27.516 | Gamma-Glutamyltransferase | U/L |
| UREA | 366654 | 0.810 | 12.390 | 5.270 | 1.630 | 5.385 | 1.282 | Urea | mmol/L |
| phenoAge | 366654 | 17.976 | 93.185 | 49.558 | 14.063 | 49.093 | 9.590 | Phenotypic Age | years |
| phenoAA | 366654 | -25.513 | 28.691 | -7.876 | 6.230 | -7.435 | 4.974 | Phenotypic Age Acceleration | years |
| ln_ALT | 366654 | 1.102 | 4.548 | 2.997 | 0.565 | 3.029 | 0.436 | Alanine Aminotransferase | ln U/L |
| ln_ALP | 366654 | 2.241 | 5.374 | 4.381 | 0.350 | 4.378 | 0.269 | Alkaline Phospatase | ln U/L |
| ln_CRP | 366654 | -2.526 | 3.195 | 0.262 | 1.413 | 0.298 | 1.024 | High Sensitivity C-Reactive Protein (CRP) | ln mg/L |
| ln_TRIG | 366654 | -1.355 | 1.929 | 0.391 | 0.712 | 0.411 | 0.505 | Triglyceride | ln mmol/L |

*ln_: marker value after natural logarithm transformation, iqr: interquartile range, Phenotypic Age: phenoAge calculated in accordance with Liu et al (Liu et al. 2018) . Phenotypic Age Acceleration: the excess of Phenotypic Age over chronological age.*

**Supplementary Table 2**: Marker key and units

| marker | name | unit |
|---|---|---|
| <chr> | <chr> | <chr> |
| phenoAge | Phenotypic Age | years |
| phenoAA | Phenotypic Age Acceleration | years |
| age | age | years |
| sex | sex | 0=female, 1=male |
| event | Dead | 0=survived, 1 = Died |
| survival_yrs | Survival time | years |
| ALB | Albumin | g/L |
| ALP | Alkaline Phospatase | U/L |
| ALT | Alanine Aminotransferase | U/L |
| APOA1 | Apolipoprotein A1 | g/L |
| APOB | Apolipoprotein B | g/L |
| CA | Calcium | mmol/L |
| CHO | Cholesterol | mmol/L |
| CREAT | Creatinine | µmol/L |
| CRP | High Sensitivity C-Reactive Protein (CRP) | mg/L |
| CYSC | Cystatin – C | mg/L |
| GGT | Gamma-Glutamyltransferase | U/L |
| GLU | Glucose | mmol/L |
| HBA1C | Glycated haemoglobin | mmol/mol |
| HDL | High Density Lipoprotein (HDL) | mmol/L |
| HLRP | High Light scatter Reticulocytes % | % |
| LDL | Low Density Lipoprotein (LDL) | mmol/L |
| LYMP | Lymphocyte percent | % |
| MCV | Mean Corpuscular (erythrocyte) Volume | fL |
| MSCV | Mean Sphered Cell Volume | fL |
| PCT | Platelet crit | % |
| PDW | Platelet Distribution Width | % |
| RDW | Red Cell (erythrocyte volume) Distribution Width | % |
| TRIG | Triglyceride | mmol/L |
| UREA | Urea | mmol/L |
| WBC | White Blood Cell or leukocyte count | x10^9 cells/L |

Supplementary Table 3 - Markers Available for each cohort/panel

| marker | age | ORCADES | Korcula | Vis | phenoAge | phenoAge' |
|---|---|---|---|---|---|---|
| <chr> | <lgl> | <lgl> | <lgl> | <lgl> | <lgl> | <lgl> |
| age | TRUE | TRUE | TRUE | TRUE | TRUE | TRUE |
| ALT | FALSE | TRUE | FALSE | FALSE | FALSE | FALSE |
| ALB | FALSE | TRUE | TRUE | TRUE | FALSE | TRUE |
| ALP | FALSE | FALSE | FALSE | FALSE | FALSE | TRUE |
| APOA1 | FALSE | TRUE | FALSE | FALSE | FALSE | FALSE |
| APOB | FALSE | TRUE | FALSE | FALSE | FALSE | FALSE |
| CA | FALSE | TRUE | FALSE | FALSE | FALSE | FALSE |
| CHO | FALSE | TRUE | TRUE | TRUE | FALSE | FALSE |
| CREAT | FALSE | TRUE | TRUE | TRUE | FALSE | TRUE |
| CRP | FALSE | TRUE | FALSE | FALSE | FALSE | TRUE |
| GGT | FALSE | TRUE | FALSE | FALSE | FALSE | FALSE |
| GLU | FALSE | TRUE | TRUE | TRUE | FALSE | TRUE |
| HBA1C | FALSE | TRUE | TRUE | TRUE | FALSE | FALSE |
| HDL | FALSE | TRUE | TRUE | FALSE | FALSE | FALSE |
| LDL | FALSE | TRUE | TRUE | FALSE | FALSE | FALSE |
| LYMP | FALSE | FALSE | FALSE | FALSE | FALSE | TRUE |
| MCV | FALSE | FALSE | FALSE | FALSE | FALSE | TRUE |
| RDW | FALSE | FALSE | FALSE | FALSE | FALSE | TRUE |
| TRIG | FALSE | TRUE | TRUE | FALSE | FALSE | FALSE |
| UREA | FALSE | TRUE | FALSE | FALSE | FALSE | FALSE |
| WBC | FALSE | FALSE | FALSE | FALSE | FALSE | TRUE |
| phenoAge | FALSE | FALSE | FALSE | FALSE | TRUE | FALSE |

*Marker codes are described in Supplementary Table 2. Each column represents one of the panels analysed. TRUE means that marker is present in the panel concerned. Note here we list phenoAge as a marker in itself, having precalculated it, and thus do not show its constituents (they are of course those of phenoAge'). PhenoAge' is a BA based on the same constituent markers as phenoAge but with coefficients recalculated to be most predictive in UKB. Marker: the code used for the marker in our analyses.*

**Supplementary Table 4a**: PlasmaAgeKorcula model (trained in UKB)

Korcula

A data.frame: 7 × 6

| marker | coeff | se | p | coeff_per_sd | uni_effect_psd |
|---|---|---|---|---|---|
| <chr> | <dbl> | <dbl> | <chr> | <dbl> | <dbl> |
| age | 0.92 | 0.01 | 0.0e+00 | 7.41 | 8.09 |
| HDL | -3.53 | 0.20 | 3.1e-68 | -1.30 | -1.99 |
| HBA1C | 0.25 | 0.01 | 2.5e-81 | 1.25 | 2.76 |
| ALB | -0.40 | 0.03 | 4.0e-52 | -1.04 | -2.21 |
| LDL | -1.18 | 0.08 | 2.0e-51 | -1.01 | -1.76 |
| CREAT | 0.06 | 0.00 | 1.6e-39 | 0.84 | 1.96 |
| GLU | 0.18 | 0.08 | 2.1e-02 | 0.15 | 1.71 |

**Supplementary Table 4b**: PlasmaAgeVis model (trained in UKB)

Vis

A data.frame: 6 × 6

| marker | coeff | se | p | coeff_per_sd | uni_effect_psd |
|---|---|---|---|---|---|
| <chr> | <dbl> | <dbl> | <chr> | <dbl> | <dbl> |
| age | 0.91 | 0.01 | 0.0e+00 | 7.34 | 8.09 |
| HBA1C | 0.27 | 0.01 | 3.9e-99 | 1.37 | 2.76 |
| CHO | -1.20 | 0.06 | 4.7e-89 | -1.34 | -1.99 |
| ALB | -0.43 | 0.03 | 2.1e-58 | -1.10 | -2.21 |
| CREAT | 0.07 | 0.00 | 1.7e-66 | 1.06 | 1.96 |
| GLU | 0.19 | 0.08 | 1.6e-02 | 0.15 | 1.71 |

*Marker: the assay code (ordered by the absolute value of effect size, measured on the SD scale). The code, assay description and units are listed in supplementary table 2.*
*Coeff: the effect in years on BA, per unit of change in the marker.*
*SE - the standard error of coeff. P- statistical significance value (two sided t-test).*
*Coeff_per_sd - change in age per one standard deviation of the predictor in the multivariate model.*
*Uni_effect_psd - change in age per one standard deviation in a univariate model.*

Supplementary Table 5: Summary statistics in ORCADES participants post QC for age, sex and biomarkers used in PlasmaAge and GlycansAge

| cohort | variable | n | min | max | median | q1 | q3 | iqr | mad | mean | sd | se | ci |
|---|---|---|---|---|---|---|---|---|---|---|---|---|---|
| <chr> | <chr> | <int> | <dbl> | <dbl> | <dbl> | <dbl> | <dbl> | <dbl> | <dbl> | <dbl> | <dbl> | <dbl> | <dbl> |
| ORCADES | sex | 595 | 0.000 | 1.000 | 0.000 | 0.000 | 1.000 | 1.000 | 0.000 | 0.452 | 0.498 | 0.020 | 0.040 |
| ORCADES | ALT | 595 | 1.000 | 190.000 | 22.000 | 17.000 | 30.000 | 13.000 | 8.896 | 26.454 | 17.520 | 0.718 | 1.411 |
| ORCADES | CRP | 595 | 0.100 | 27.800 | 1.180 | 0.591 | 2.465 | 1.873 | 1.072 | 2.183 | 3.060 | 0.125 | 0.246 |
| ORCADES | age | 595 | 40.000 | 70.000 | 56.200 | 47.450 | 62.550 | 15.100 | 10.971 | 55.236 | 8.716 | 0.357 | 0.702 |
| ORCADES | ln_ALT | 595 | 0.000 | 5.247 | 3.091 | 2.833 | 3.401 | 0.568 | 0.382 | 3.145 | 0.485 | 0.020 | 0.039 |
| ORCADES | ALB | 595 | 28.000 | 52.000 | 42.000 | 40.000 | 44.000 | 4.000 | 2.965 | 41.780 | 3.154 | 0.129 | 0.254 |
| ORCADES | APOA1 | 595 | 0.060 | 3.362 | 0.920 | 0.781 | 1.104 | 0.323 | 0.228 | 0.988 | 0.357 | 0.015 | 0.029 |
| ORCADES | APOB | 595 | 0.009 | 2.256 | 0.512 | 0.313 | 0.765 | 0.453 | 0.314 | 0.566 | 0.322 | 0.013 | 0.026 |
| ORCADES | CREAT | 595 | 39.000 | 230.000 | 78.000 | 69.000 | 88.000 | 19.000 | 14.826 | 80.039 | 16.896 | 0.693 | 1.360 |
| ORCADES | ln_CRP | 595 | -2.303 | 3.325 | 0.166 | -0.525 | 0.902 | 1.427 | 1.043 | 0.212 | 1.050 | 0.043 | 0.085 |
| ORCADES | GGT | 595 | 5.000 | 428.000 | 22.000 | 15.000 | 36.000 | 21.000 | 11.861 | 32.570 | 35.498 | 1.455 | 2.858 |
| ORCADES | GLU | 595 | 4.000 | 15.500 | 5.300 | 5.000 | 5.700 | 0.700 | 0.445 | 5.510 | 0.984 | 0.040 | 0.079 |
| ORCADES | HBA1C | 595 | 21.000 | 97.000 | 36.000 | 33.000 | 39.000 | 6.000 | 4.448 | 37.029 | 8.427 | 0.345 | 0.679 |
| ORCADES | HDL | 595 | 0.700 | 3.320 | 1.500 | 1.280 | 1.800 | 0.520 | 0.385 | 1.560 | 0.418 | 0.017 | 0.034 |
| ORCADES | LDL | 595 | 0.000 | 9.000 | 3.000 | 3.000 | 4.000 | 1.000 | 1.483 | 3.576 | 1.080 | 0.044 | 0.087 |
| ORCADES | UREA | 595 | 2.664 | 18.981 | 5.495 | 4.829 | 6.494 | 1.665 | 1.234 | 5.764 | 1.549 | 0.064 | 0.125 |
| ORCADES | PlasmaAA | 595 | -19.309 | 25.384 | -0.671 | -4.827 | 3.203 | 8.031 | 6.052 | -0.600 | 6.516 | 0.267 | 0.525 |
| ORCADES | PlasmaAge | 595 | 23.056 | 92.433 | 54.436 | 46.163 | 62.585 | 16.423 | 12.185 | 54.636 | 11.069 | 0.454 | 0.891 |
| ORCADES | GP6 | 595 | 1.749 | 10.443 | 4.520 | 3.700 | 5.581 | 1.881 | 1.360 | 4.743 | 1.472 | 0.060 | 0.119 |
| ORCADES | GP14 | 595 | 4.328 | 27.370 | 14.187 | 11.984 | 16.278 | 4.294 | 3.171 | 14.331 | 3.420 | 0.140 | 0.275 |
| ORCADES | GP15 | 595 | 0.742 | 3.116 | 1.561 | 1.330 | 1.821 | 0.491 | 0.362 | 1.598 | 0.368 | 0.015 | 0.030 |
| ORCADES | GlycanAge | 595 | 25.474 | 79.281 | 54.880 | 48.258 | 61.117 | 12.859 | 9.560 | 54.581 | 9.597 | 0.393 | 0.773 |
| ORCADES | GlycanAA | 595 | -25.956 | 24.322 | -0.809 | -6.057 | 4.971 | 11.028 | 8.493 | -0.657 | 8.213 | 0.337 | 0.661 |
| ORCADES | GlycanA'A | 595 | -26.245 | 21.383 | 0.014 | -5.090 | 4.929 | 10.020 | 7.360 | 0.000 | 7.666 | 0.314 | 0.617 |
| ORCADES | GlycanAge' | 595 | 22.940 | 82.814 | 56.004 | 46.957 | 64.097 | 17.139 | 12.834 | 55.237 | 11.610 | 0.476 | 0.935 |

*Marker codes and units are described in Supplementary Table 2.*

**Supplementary Figure 1a**: The distribution of blood biomarker in UKBB - 4 markers alanine transferase, alkaline phosphatase, C-reactive protein and triglycerides (ALT ALP CRP and TRIG) show substantial right skew

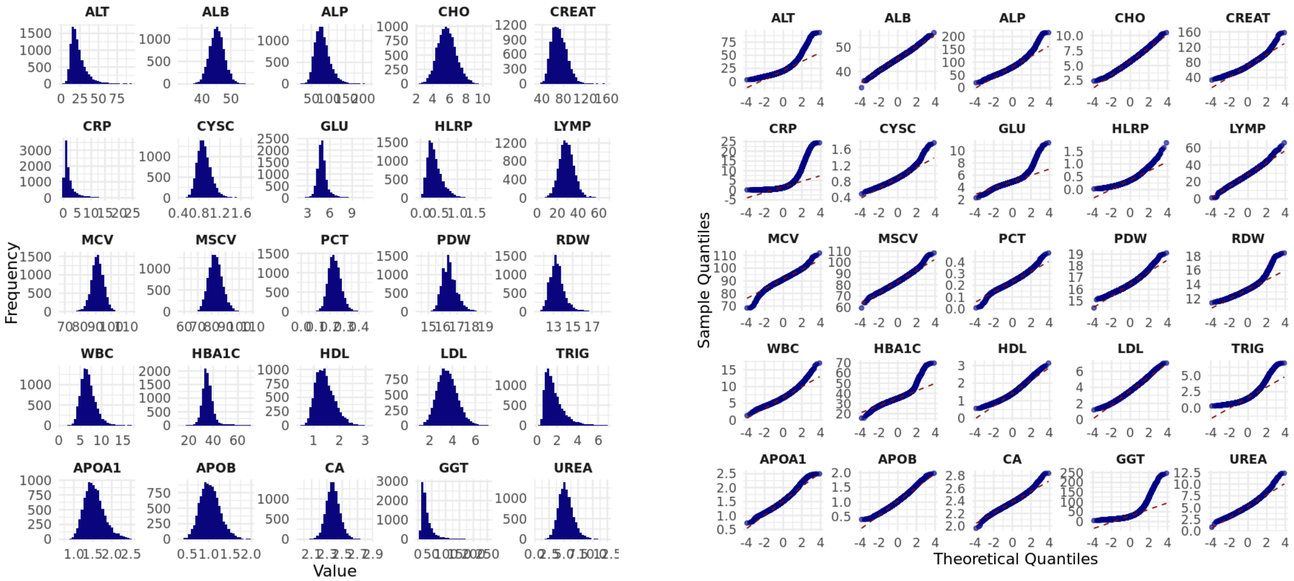


*The key to marker codes is in Supplementary Table 2. The second panel is the q-q plot against the normal distribution.*

**Supplementary Figure 1b**: The distribution in UKBB of alanine transferase, alkaline phosphatase, C-reactive protein and triglycerides are less skewed after log transformation.

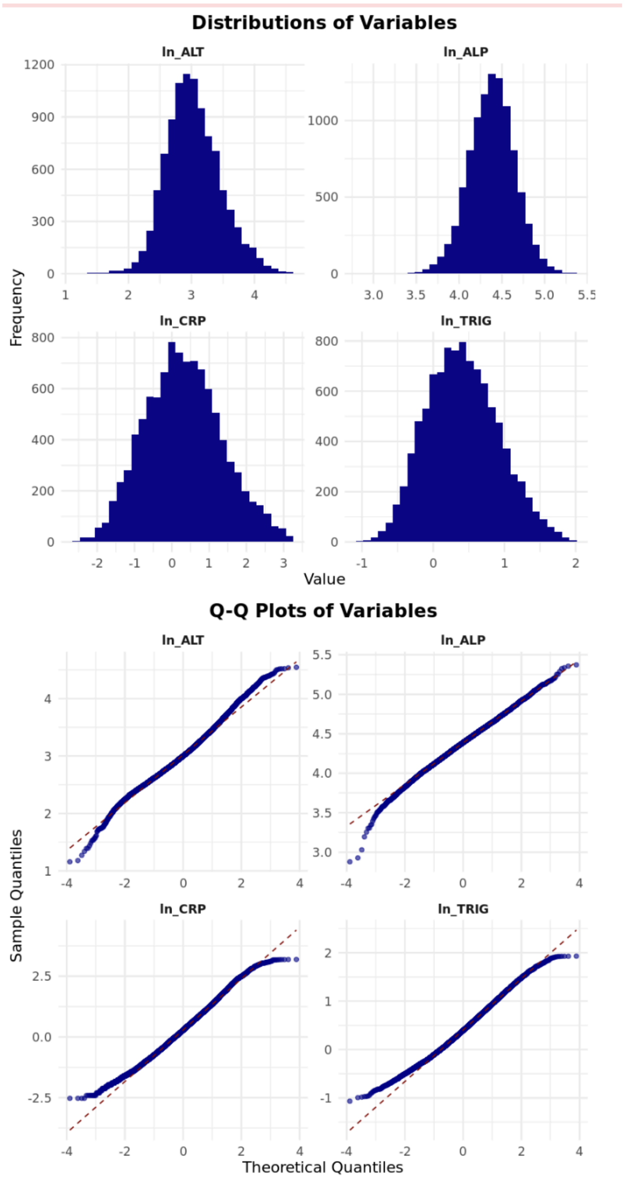


*The key to marker codes is supplementary table 2. ln_: marker value after natural logarithm transformation. The second panel is the q-q plot against the normal distribution.*

Supplementary Figure 2 - Waterfall Chart of ORCADES QC

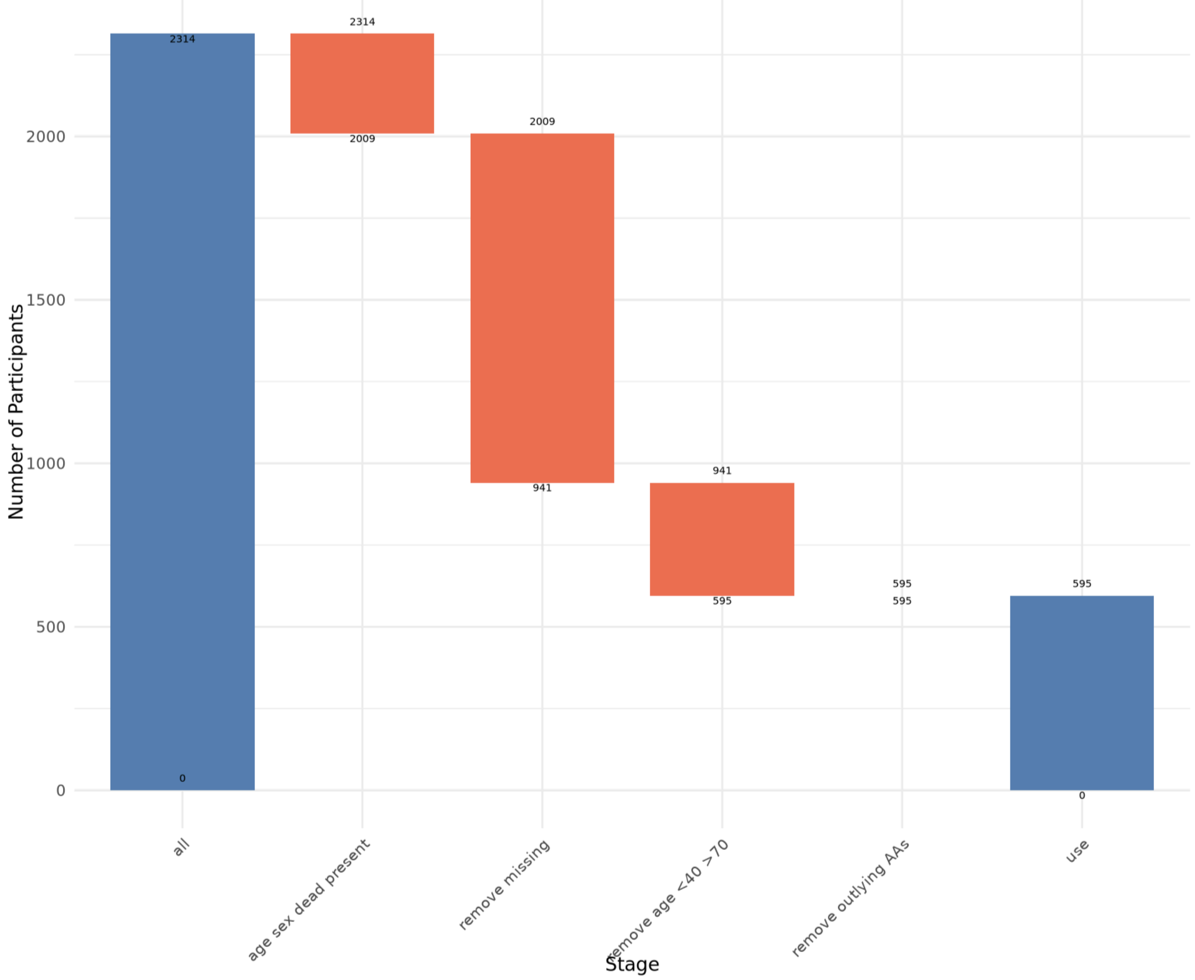

**Supplementary Fig. 3:** Glycan Age is an unbiased predictor of chronological age in ORCADES

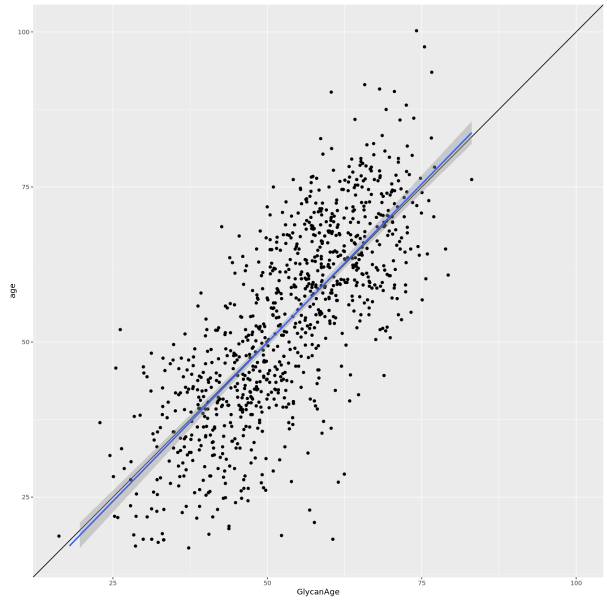


*Participants with a GlycanAge x, have, broadly speaking, a mean chronological age of x. Note to aid clarity of the point being made, this figure shows participants pre-qc (in particular before the age restriction).*

**Supplementary Fig. 4: For participants with a given higher and lower age, mean GlycanAge is not equal to chronological age and GlycansAA is thus (undesirably for our purposes) correlated with age.**

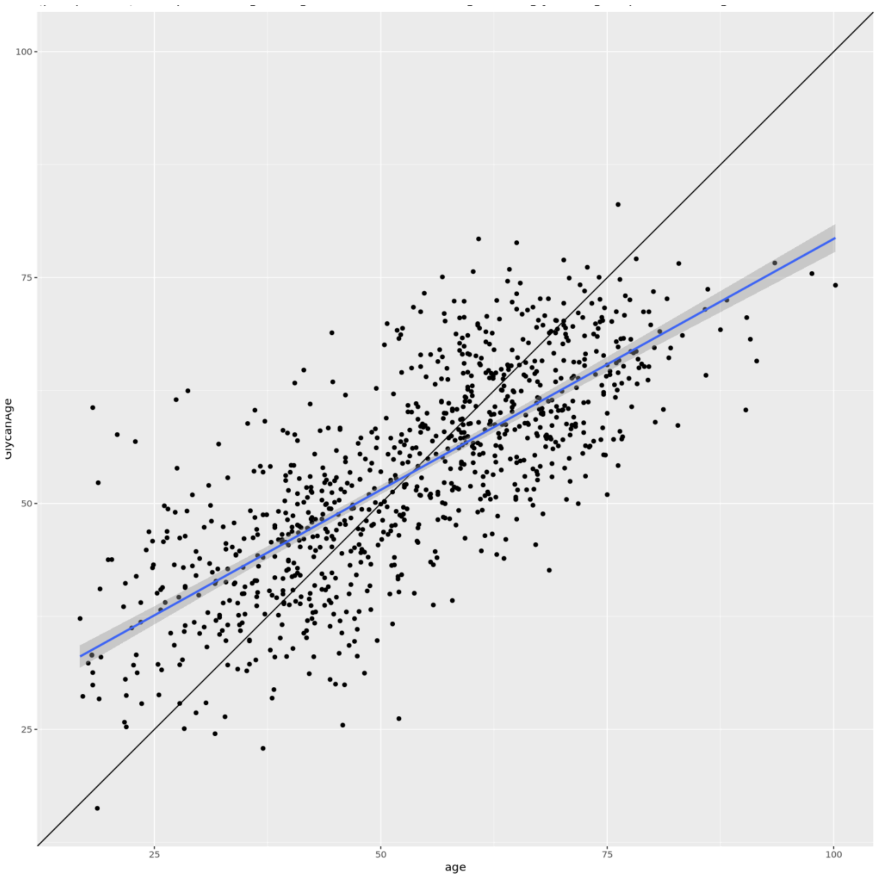


*Participants aged for example 25 appear to have a mean GlycanAge ~ 37, and conversely participants aged 75 appear to have a mean GlycanAge ~ 62. Note to aid clarity of the point being made, this figure shows participants pre-qc (in particular before the age restriction).*

**Supplementary Fig. 5**: Participants at a given age, have a mean GlycanAge' equal to their age and GlycanA'A is not correlated with chronAge.

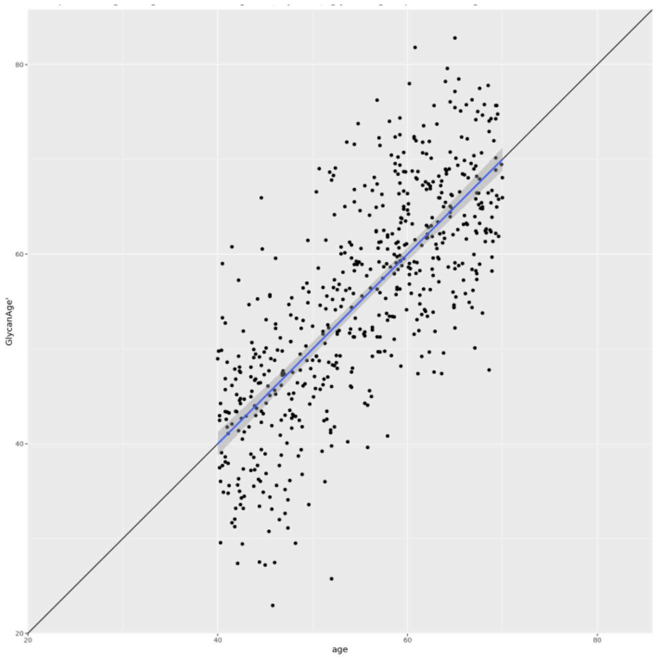


*age: chronological Age*
*GlycanAge', recalibrated GlycanAge so there is no correlation between glycansAA' and chronological age (and line of best fit is y=x).*

**Supplementary Fig. 6**: ROC curves for mortality in ORCADES in models with Chronological age and Biological Ages.

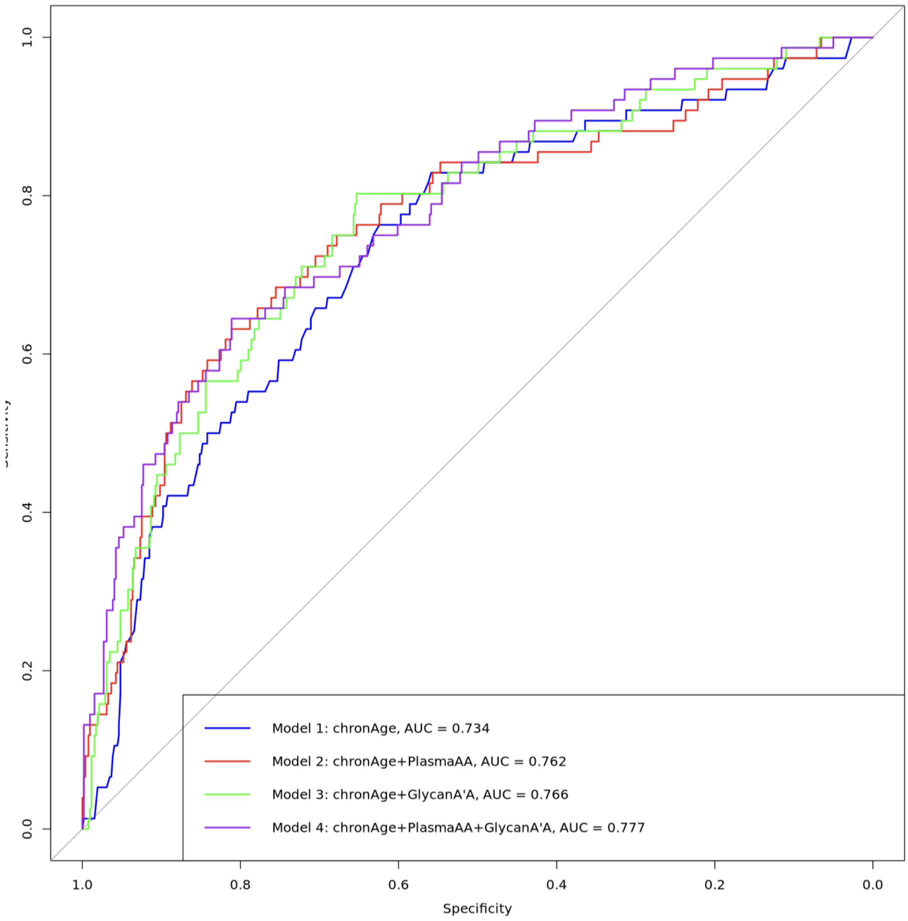

**Supplementary Fig. 7**: Forest plots representing meta-analysis of AA effects on survival in three Croatian cohorts. a) chronAge + PlasmaAA model b) chronAge + GlycansAA model c) chronAge + PlasmaAA + GlycansAA model. The effect and its 95% CI are reported as black horizontal lines for individual cohorts. Red line represents overall effect and 95% CI.

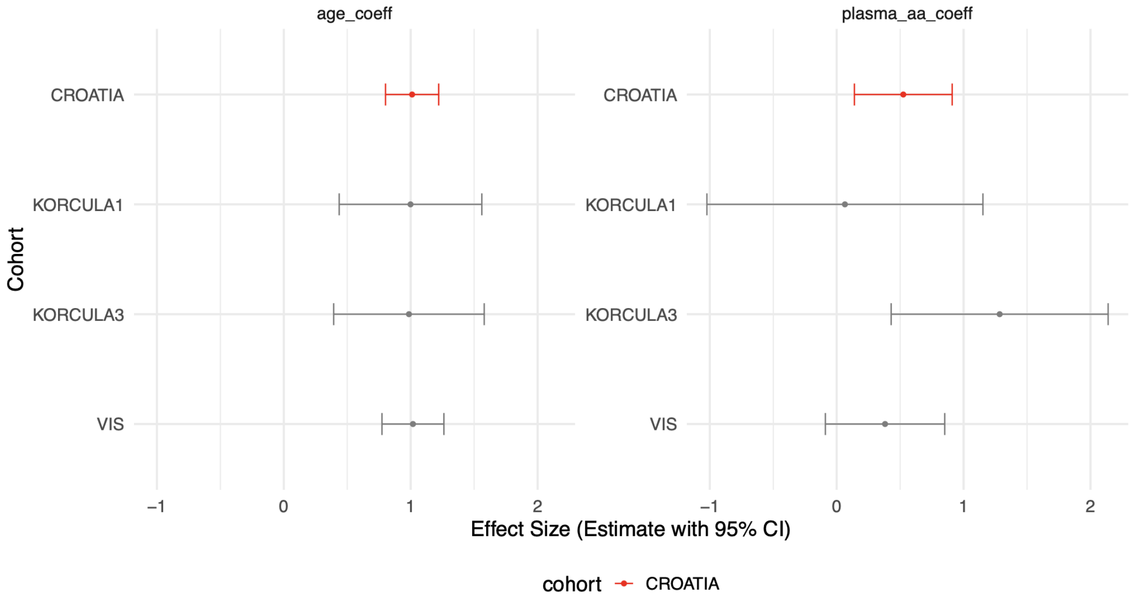


b)

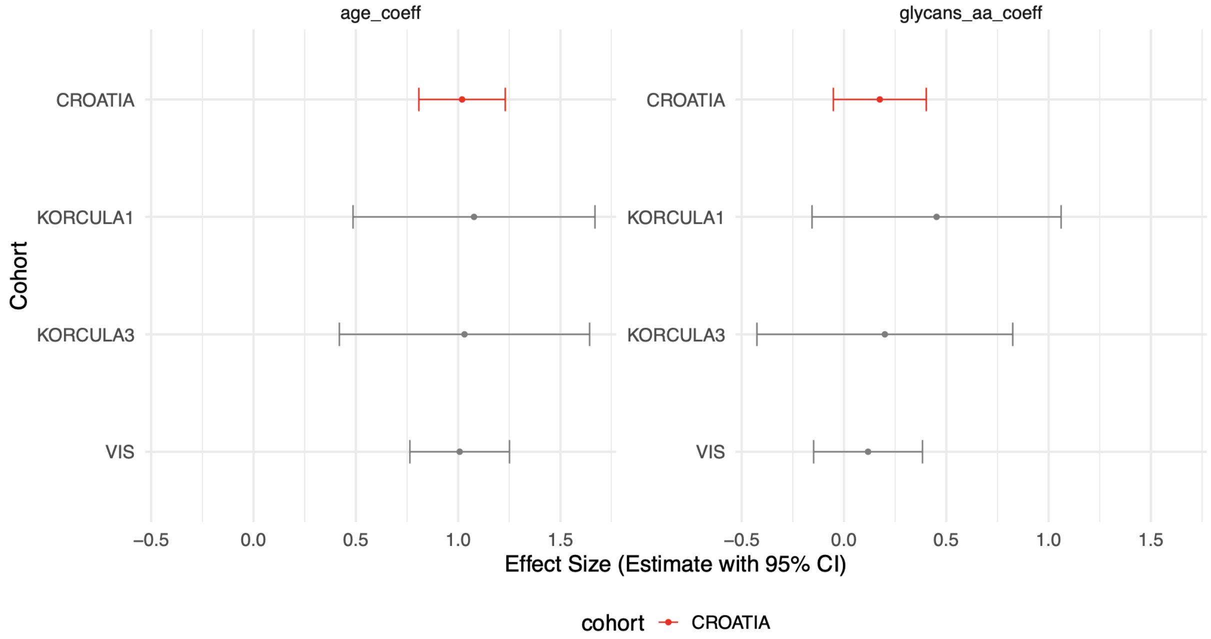

c)

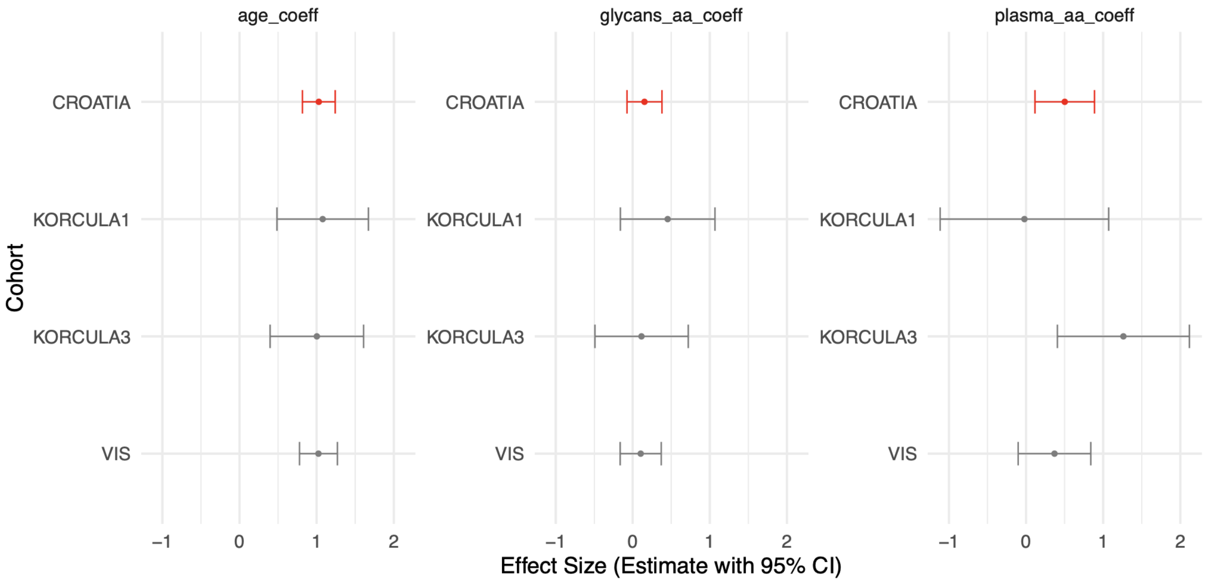


**Supplementary Fig. 8**: ROC curves for tested models for mortality prediction in CROATIA cohorts. a) Vis, b) Korcula3, c) Korcula1

a) b) c)

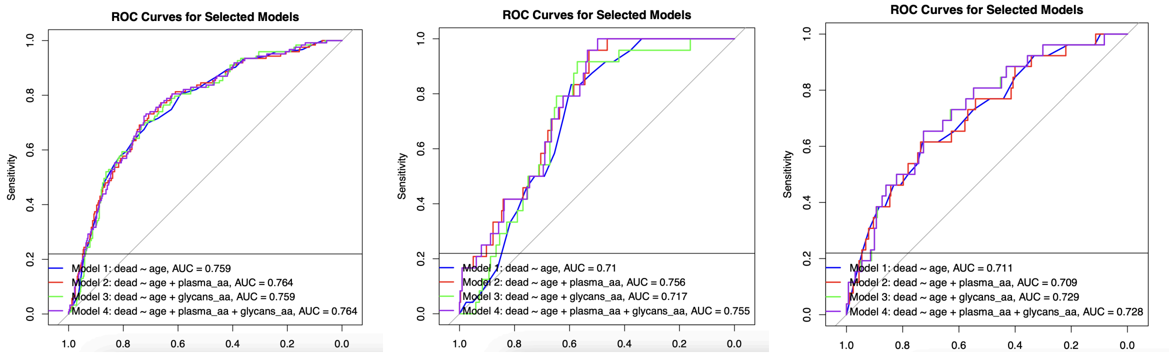
